# Familial factors inflate the estimated effects of genetic liability to ADHD on behavioural outcomes

**DOI:** 10.64898/2026.10.27.26366008

**Authors:** Daisy C. P. Crick, Neil M. Davies, Sarah E. Medland, David M. Evans

**Affiliations:** Division of Psychiatry, University College London, London WC1E 6BT, UK; Institute for Molecular Bioscience, University of Queensland, Brisbane, Australia; Department of Statistical Science, University College London, London WC1E 6BT, UK; Department of Public Health and Nursing, Norwegian University of Science and Technology, Norway; Psychiatric Genetics, QIMR Berghofer, Brisbane, Australia; The Frazer Institute, The University of Queensland, Woolloongabba, QLD, Australia; MRC Integrative Epidemiology Unit, University of Bristol, Bristol, UK

## Abstract

**Background:** People with ADHD have worse health, behavioural and social outcomes, widely interpreted as evidence that ADHD causes these harms, justifying early identification and treatment. But ADHD clusters in families, and conventional studies cannot separate the effect of a person’s own ADHD from the family environment in which it arises. We used a family-based genetic design to distinguish the two.

**Methods:** We used up to 22 genome-wide significant variants associated with ADHD (38,691 cases, 186,843 controls) to estimate effects of genetic liability to ADHD on 16 health and behavioural outcomes. We compared population-based estimates with family-based estimates from within-sibship analyses of up to 164,220 siblings. We tested for heterogeneity between the designs, and applied sensitivity analyses. We involved individuals with lived experience of ADHD in interpreting the findings and shaping how the clinical/public-health implications were described.

**Findings:** Population-based analyses implied that greater genetic liability to ADHD increased smoking, alcohol use and depressive symptoms, and reduced educational attainment, age at first child and cognitive performance. In family-based analyses, these effects attenuated. For example, the effect on number of depressive symptoms fell from 0·059 SD (95% CI 0·039, 0·080) to 0·002 SD (–0·052, 0·055), and age at first child increased from –0·045 SD (–0·058, –0·033) to 0·003 SD (–0·040, 0·047), with significant differences between designs (p=0·048 and p=0·034). Findings were consistent across sensitivity analyses.

**Interpretation:** Much of the excess health and behavioural burden attributed to ADHD may reflect the family and environmental context in which ADHD occurs, rather than an individual’s own ADHD. Interventions targeting the individual might not reduce these outcomes as much as expected. Distinguishing such effects requires family-based designs.

**Funding:** Medical Research Council, UK Research and Innovation, Research Council of Norway, US National Institute of Mental Health, and Australian National Health and Medical Research Council.

**Research in context:** *Evidence before this study:* Attention-deficit hyperactivity disorder (ADHD), and higher genetic liability to ADHD, are associated with a wide range of adverse health, behavioural, and social outcomes. A 2024 umbrella review of the impacts of ADHD summarised consistent associations with addiction, mental and physical illness, lower educational attainment, and social difficulties. Earlier reviews of Mendelian randomization studies of ADHD reached similar conclusions, reporting apparent causal effects on smoking, depression, body-mass index, and educational attainment. A broad two-sample Mendelian randomization analysis of ADHD across more than 100 traits corroborated these findings. Almost all of this genetic evidence, however, comes from samples of unrelated individuals, in which estimates can be confounded by population stratification, assortative mating, and dynastic effects. We searched PubMed and Google Scholar from database inception to 20/07/26 using terms related to ADHD, Mendelian randomization, and within-family designs (**Supplementary note**), without language restrictions. We identified eight relevant studies, however only three used a within-family or trio Mendelian randomization design. None used a large within-family Mendelian randomization design to investigate the effects of genetic liability to ADHD on health or behavioural outcomes while separating individual’s liability from familial influences.

*Added value of this study:* To our knowledge, this is the largest within-family Mendelian randomization study of genetic liability to ADHD, comparing conventional population-based estimates with family-based estimates across 16 outcomes in up to 164,220 siblings. Population-based estimates reproduced previously reported associations, but most attenuated in family-based analyses. For depressive symptoms and age at first child, the two designs significantly differed, indicating that the population-based estimates were inflated by familial factors.

*Implications of all the available evidence:* Much of the excess health, behavioural, and social burden attributed to ADHD may reflect the family and environmental context in which ADHD arises, rather than an individual’s own liability. If so, interventions directed solely at the individual may not reduce these outcomes as much as observational and population-based estimates imply. These findings should be interpreted alongside the broader evidence rather than as a basis for immediate changes in practice, and they underscore the need for larger family-based genetic studies.

---

Attention-deficit hyperactivity disorder (ADHD) is a neurodevelopmental disorder characterised by age-inappropriate impulsivity, hyperactivity, and inattention^[1, 2]^. Twin and family-based studies consistently estimate the heritability of ADHD to be ∼70-80% across populations, with males more likely to be diagnosed with ADHD than females (∼10 vs 5%)^[3]^. Individuals diagnosed with ADHD typically have poorer health outcomes compared to controls. Whether these adverse outcomes are caused by ADHD itself, and are therefore potentially modifiable by treating the individual, or reflect the family and social environments in which ADHD arises, has not been established. Yet, this distinction determines where intervention should be targeted.

In 2024, an umbrella review identified three key domains of risk associated with an ADHD diagnosis: mental health, physical health, and social/behavioural factors^[4]^. The findings suggested that ADHD diagnoses were associated with increased rates of addiction, mental health disorders, obesity, type-2 diabetes mellitus, criminality, lower educational attainment, relationship difficulties and unplanned pregnancies. Similar findings have been reported previously^[5–11]^ and associations have also been found with early mortality^[12]^. However, associations reported in conventional observational epidemiological studies are susceptible to confounding^[13]^ and may not reflect a causal effect of ADHD on subsequent health-related outcomes.

Mendelian randomization^[14–16]^ is an epidemiological method that uses genetic variants as instrumental variables (IVs) by exploiting the random transmission of genetic variation from parents to their offspring. When used appropriately, Mendelian randomization can potentially overcome unmeasured confounding of the exposure-outcome relationship^[17]^. For causal inference from Mendelian randomization studies to be valid, the genetic variant(s) must meet three core IV assumptions (**Figure 1**).^[15, 18–20]^.

**Figure 1:**
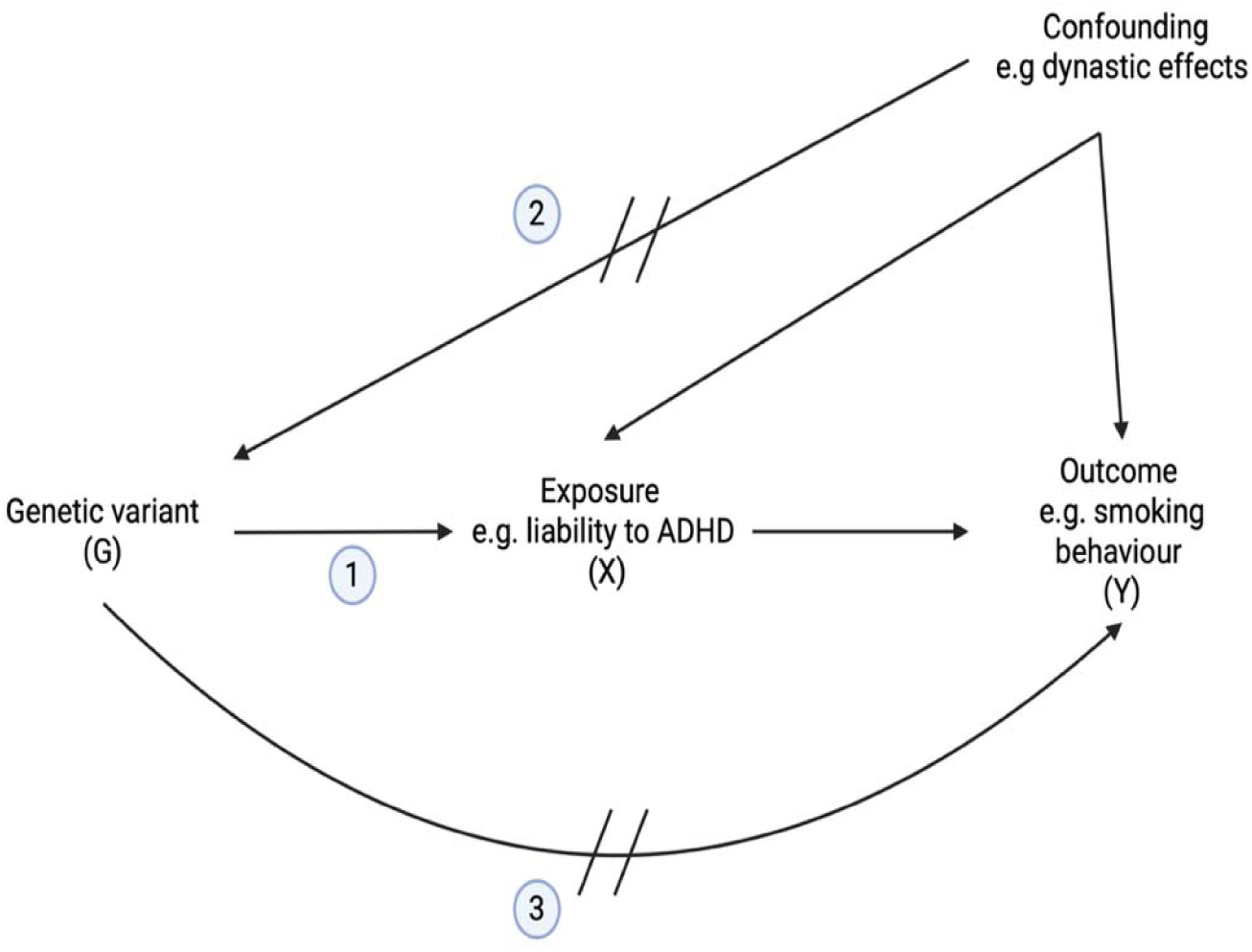
Direct Acyclic Diagram illustrating the core assumptions necessary for using a genetic variant as an instrumental variable for causal inference in Mendelian randomization studies. *Assumption 1 (Relevance): The instrument (G) is robustly associated with the exposure (X).* *Assumption 2 (Independence): There are no uncontrolled confounders of the association between the instruments and the outcome, i.e. the instrument does not have a common cause with the outcome.* *Assumption 3 (Exclusion-restriction): The instrument influences the outcome (Y) only through its effect on the exposure*.

A review of 47 Mendelian randomization studies suggested that those with higher genetic liability to ADHD were more likely to start smoking, develop depression, have higher BMI and lower educational attainment and income^[21]^. Many of these findings were replicated in a subsequent two-sample Mendelian randomization study that investigated the relationship between ADHD and 124 traits^[22]^. The results suggest genetic liability to ADHD may have a causal effect on health-related behaviours/outcomes across the lifespan (key results from this literature summarised in **eTable 1**^[21–24]^). However, the majority, if not all, of the Mendelian randomization studies investigating the effects of ADHD on health to date have used samples of unrelated individuals, which we will refer to as population-based Mendelian randomization. Given that Mendelian segregation randomizes genetic variants *within families*^[25]^, population-based Mendelian randomization estimates may be inconsistent if genetic variants associated with ADHD are not randomly distributed across the population^[26–29]^ (i.e. there is uncontrolled confounding of the genetic variant -outcome association). This is because it would violate the independence assumption.

Confounding of the genetic variant-outcome association can occur through environmental and social factors, including population stratification, assortative mating (i.e. selection of mates on the basis of trait similarity) and/or dynastic effects (i.e. genetic nurture) whereby expression of parental genotype in the parental phenotype in turn affects the offspring phenotype^[29]^. These mechanisms may create spurious nondirect associations between genetic variants and outcomes, leading to inconsistent estimates of the variants’ causal effects in offspring (**Figure 2**)^[26]^. This is important to consider given that ADHD may be under phenotypic assortment^[30, 31]^ and dynastic effects are likely to be relevant (**Box 1**)^[22]^.

**Figure 2:**
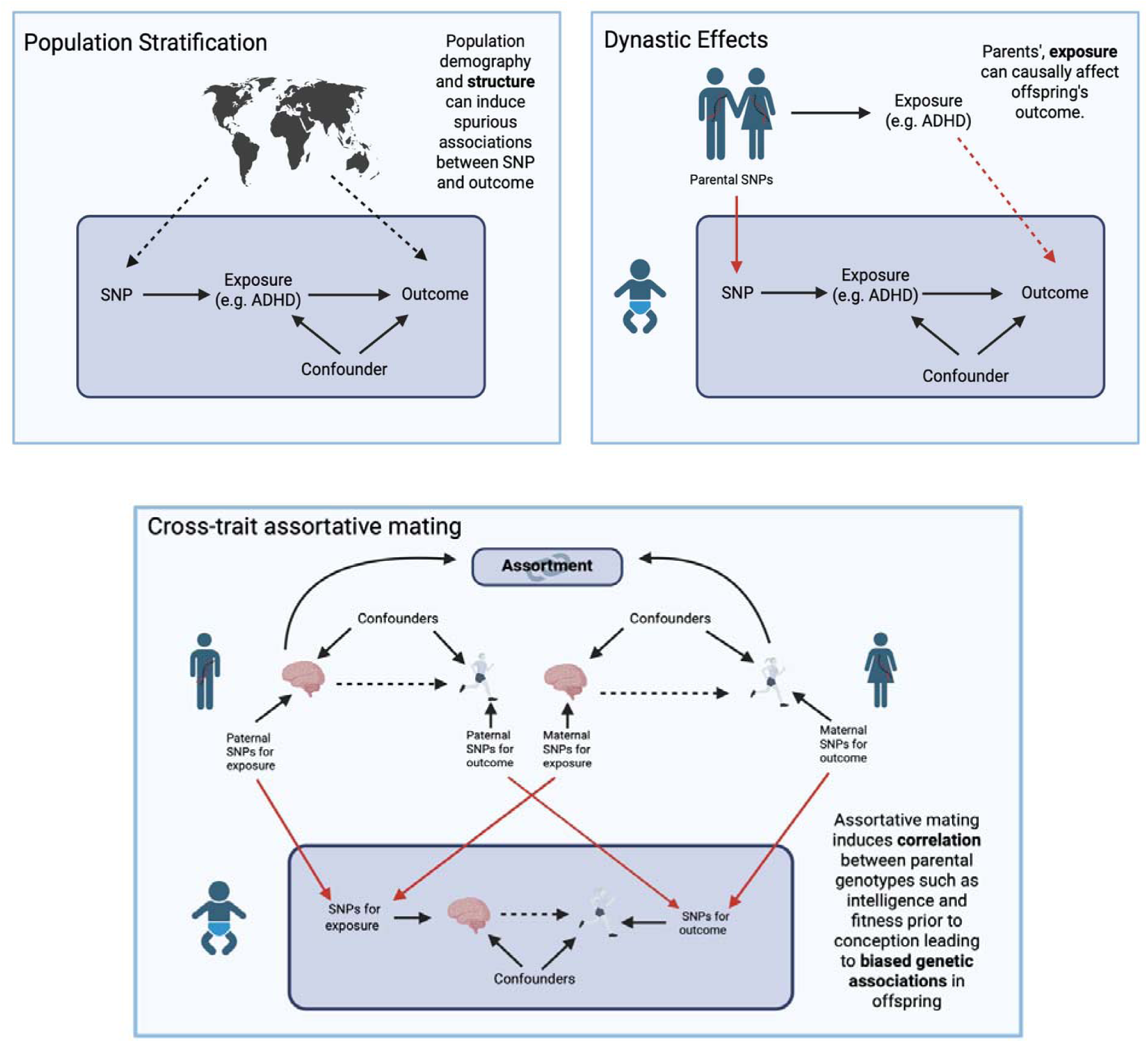
Schematic representation of the confounding effect of population stratification,cross-trait assortative mating and dynastic effects in population based Mendelian randomization studies (adapted from Brumpton et al 2020^[26]^). Solid arrows denote causal paths, dashed arrows represent confounded associations. Red arrows depict genetic transmission from parents to offspring. Mendelian randomization estimates of the causal effect of the exposure on the outcome can be inconsistent because of back door paths between the exposure SNPs and the outcome. **Population stratification**: The presence of different genetic ancestry groups within the same genetic association study. It induces spurious associations between genetic variants and phenotypes through shared ancestry rather than biological causation. **Cross-trait assortative mating:** Males who are high on the exposure variable are more likely to mate with females who are high on the outcome variable. This generates a confounding path between the SNPs for the exposure in the offspring generation and the outcome variable in this generation. **Dynastic effects**: Individuals who have ADHD are more likely to have parents who also have ADHD. Offspring outcomes can be influenced indirectly via the environment that the parent provides, violating the independence assumption

In contrast, within-family Mendelian randomization studies, which use genotyped sibling pairs or parent-offspring trios, can control for the offspring’s familial genetic background at the molecular genetic level^[25]^. Within-family studies can either use genotypic differences between siblings or condition on parental genotypes (parent-offspring trios). By exploiting the consequences of random Mendelian segregation within families, both approaches are robust to population stratification, assortative mating and dynastic effects^[27, 28]^.

In the present study, we compared two-sample population-based and within-family Mendelian randomization estimates of the effect of genetic liability to ADHD on health, behavioural and social outcomes. Given that within-family designs separate a person’s own genetic liability from the familial and environmental factors that confound conventional genetic studies, this tests whether the outcomes associated with ADHD reflect the individual’s own ADHD. To our knowledge, this is the largest molecular genetic family-based study of ADHD to date.

## Box 1

### Sources of inconsistency in population-based Mendelian randomization studies

#### Example of population stratification

Population stratification refers to the presence of different genetic ancestry groups within the same genetic association study^[32]^. When individuals from different ancestries differ in both allele frequencies and disease prevalence (or mean trait value), genetic variants can appear to be associated with the disease or trait, but only because ancestry is correlated with both. Consequently, this can lead to spurious associations or, conversely, obscure genuine associations. A classic example of population stratification involves a genetic variant near the promoter of the LCT gene, which influences the persistence of lactase production into adulthood^[33]^. The frequency of this variant shows a strong latitudinal gradient across Europe. Consequently, the variant may appear to be associated with traits that also vary geographically across this gradient, even when there is no direct causal relationship between the variant and the trait.

Population stratification can be controlled effectively by using principal components, although residual confounding due to more recent or fine-scale population structure may remain. Additionally, within-family tests of genetic association are robust to population stratification because they compare genetic differences between relatives, thereby controlling for differences in ancestry between families.

#### Example of cross-trait assortative mating.

Cross-trait assortment is the phenomenon where mates display cross-correlations across distinct traits^[34]^. Cross-trait assortment on two traits can lead parents to be genetically correlated on traits. This assortment means that the traits are likely to be genetically correlated in the offspring which can lead to confounding in Mendelian randomization. One known example of cross-trait assortative mating, is that more educated people often demonstrate preferences for taller partners, meaning that there is a positive correlation between a person’s height and their mate’s educational attainment^[35]^. As a result, genetic variants associated with educational attainment can become correlated with variants associated with height, even if there is no direct causal effect of one trait on the other. This association can be amplified if the same patterns of assortment occur over generations. Similar processes may occur with ADHD, particularly for socially influenced outcomes (e.g. educational attainment)^[34, 36, 37]^.

#### Example of dynastic effects

Individuals who have ADHD are more likely to have parents who also have ADHD^[38, 39]^. As such, variants for ADHD in the offspring generation are also likely to be associated with parental ADHD behaviours, symptoms and diagnoses. Therefore, the effect of the “expressed” parental ADHD on offspring health can confound the causal estimate of genetic liability to ADHD on health-related outcomes in population-based Mendelian randomization studies. In other words, the population-based causal estimate will also reflect nondirect population-level phenomena operating through parental ADHD and the familial environment.

If genetic liability to ADHD increases the likelihood of smoking, pregnant people with higher liability to ADHD may be more likely to smoke during pregnancy or around birth compared to those without the genetic variants^[40, 41]^. Therefore, children who inherit more ADHD-associated variants may have had greater exposure to smoking whilst *in utero*. Given that smoking during pregnancy is associated with neurodevelopmental outcomes such as ADHD in offspring^[37, 38]^, if the variants were inherited by the child, a spurious association between the child’s smoking genotype and their ADHD symptoms could be induced, even if their own smoking behaviour plays no causal role.

## Methods

### Selection of the genetic instruments

Genome-wide significant single-nucleotide polymorphisms (SNPs) (P<5.0×10^-8^) were used as instruments for ADHD and identified from summary-level population-based data from a European GWAS sex-combined meta-analysis (38,691 individuals with ADHD and 186,843 controls)^[42]^. The GWAS reported 27 genome-wide significant SNPs; however, the maximum number of SNPs used in our analysis was 22. This was because SNPs were removed during the harmonization process (if they were palindromic with intermediate allele frequencies or during LD clumping because they were correlated with retained SNPs).

For each phenotype, we used the same set of SNPs in the population-based and family-based Mendelian randomization analyses to ensure that the estimates were directly comparable. The SNPs, their closest genes, their putative functions, and known associations are presented in **eTable 2**. Because SNP–exposure associations came from a population-based GWAS, we tested robustness to their inflation across a range of shrinkage values (**Supplementary Note**).

Using individual-level data from UK Biobank (UKB)^[43]^, we investigated whether the genetic variants were associated with maternal smoking status during pregnancy or with own birthweight. This was to assess the validity of the Mendelian randomization assumptions^[17, 44]^ (**Supplementary Note**).

### Outcome phenotypes

We used family-based (sibling) summary data from a Within Family Consortium meta-analysis GWAS comprising 18 European cohorts (independent of the exposure cohorts) and up to 164,220 siblings^[29]^. From this, 16 outcome phenotypes of interest were chosen to be investigated based on previously reported observational associations with ADHD^[5–10]^. The family-based SNP-outcome associations control for effects of population stratification, dynastic effects, and assortative mating that may confound conventional Mendelian randomisation estimates obtained from unrelated individuals.

Summary statistics for the population-based outcome estimates were identified using the Integrative Epidemiology Unit OpenGWAS platform^[45]^. We aimed to use the largest available sample size while maintaining phenotypic similarity with the Within Family Consortium phenotypes. Information about sample sizes used for each outcome and the OpenGWAS codes (https://opengwas.io) are presented in **eTable 3**. To check that between-design differences did not reflect differences in samples, we repeated the population-based analysis using Within Family Consortium summary statistics.

A table of the number of SNPs removed for being palindromic or in linkage disequilibrium (LD) for each phenotype is reported on GitHub (https://github.com/daisycrick/Within_family_ADHD).

### Statistical Analysis – Two-Sample Mendelian Randomization

We conducted two-sample Mendelian randomization analyses to estimate the effect of genetic liability to ADHD on health outcomes and to explore whether causal estimates from conventional population-based Mendelian randomization studies were likely to reflect population stratification, assortative mating and/or dynastic effects. We used the same population-based summary statistics to select exposure instruments for both approaches. However, for the conventional Mendelian randomization analysis we used population-based summary statistics (unrelated samples) for the outcomes, whereas for the family-based Mendelian randomization we used family-based (sibling-pairs) summary data for the outcomes. SNPs were harmonised to the same effect allele; palindromic, unavailable, and correlated SNPs were handled by standard procedures (outlined in the **Supplementary Note**). We calculated an F-statistic for each SNP to assess the strength of association between the instrument and ADHD^[14]^ (**Supplementary Note**).

Post-harmonisation, 21-22 SNPs were available to investigate the effect of a genetic liability to ADHD on the health and behavioural outcomes in both the population-based and family-based analyses.

We used the inverse-variance weighted (IVW) estimator^[46]^ in the primary analysis, which assumes no unbalanced directional horizontal pleiotropy. The weighted median and weighted mode methods were used in the sensitivity analyses^[47]^. Consistency in results between methods strengthens causal inference because each method makes distinct assumptions about instrument validity. In contrast, divergent results may suggest a violation of the third Mendelian randomization assumption (the exclusion restriction assumption)^[47–49]^. Outcomes were standardised using the population standard deviation (**Supplementary Note**, **eTable 3**). Mendelian randomization effect estimates and standard errors were multiplied by ln(2) (0.693), meaning that estimates are expressed as standard deviation units of the outcome per doubling of the genetically predicted odds of ADHD.

We report nominal p-values and p-values corrected for multiple testing (n=16 phenotypes) using the Benjamini–Hochberg false discovery rate (FDR) at q < 0.05.

### Sensitivity Analyses

Presence of heterogeneity in causal effect estimates across individual SNPs was assessed using Cochran’s Q test in the IVW analyses. We used the Mendelian randomization Pleiotropy Residual Sum and Outlier global test (MR-PRESSO) to investigate the presence of pleiotropy^[50]^. Finally, we formally tested the extent to which causal estimates from the population-based Mendelian randomisation analyses differed from the family-based Mendelian randomisation estimates using a Wald test^[51]^. We assumed zero covariance between designs; bivariate LDSC across contributing cohorts supported minimal correlated estimation error (**Supplementary Note**).

By using the genetic differences between siblings, the within family effect estimates are robust to population stratification, assortative mating, and dynastic effects^[27, 28]^. However, other methods control for offspring’s familial genetic background at the molecular genetic level. For example, trio analyses exploit the segregation of alleles within families while parent-offspring duo analyses condition on the genotype of one parent^[25]^. Assumptions differ across within-family designs, and each is also liable to different sources of inconsistency. We discuss the different within-family frameworks, describe their assumptions for the population, trios, and duos models, and compare them in the **Supplementary Note**.

### Patient and public involvement

We involved 10 young people with lived experience of ADHD in interpreting the findings and shaping how the clinical and public-health implications were described. Their input informed our framing of which harms may be modifiable and helped ensure that the findings were not presented as minimising ADHD or its treatment. They were not involved in the study design or analysis, which used existing summary-level and de-identified genetic data.

## Results

We investigated whether the instrumental variable assumptions were likely to hold. The mean F-statistic across the 22 ADHD-associated variants was 870.0 (651.7-1759.5; available on GitHub), indicating that the relevance assumption holds and that our analysis is unlikely to be biased by weak instruments.

To investigate potential residual confounding and violations of the second Mendelian randomization assumption (the independence assumption), we examined whether the genome-wide significant ADHD SNPs were associated with maternal smoking during pregnancy using individual-level UKB data (**eTable4**). Estimates were orientated to the ADHD-increasing allele. One ADHD-associated SNP (rs1438898) was associated with maternal smoking during pregnancy after Bonferroni correction. Maternal smoking during pregnancy precedes offspring ADHD which means that the association may reflect population stratification, assortative mating or dynastic effects. This provides further rationale for the use of family-based Mendelian randomization analysis, which is likely to be robust to confounding arising from population stratification, assortative mating and dynastic effects.

We also investigated the association between the genome-wide significant ADHD SNPs and own birthweight using individual-level UKB data (**eTable4**). Again, estimates were orientated to the ADHD-increasing allele. After Bonferroni correction, several SNPs were associated with own birthweight (rs549845, rs2311059, rs6537401, rs9969232, rs11596214, rs2582895). Although the effect direction was not consistent across SNPs, the number of associations exceeded what would be expected by chance. Here, associations could be due to horizontal pleiotropy^[17, 44]^. We therefore considered these findings alongside the pleiotropy-robust analyses.

The population-based Mendelian randomization analysis replicated previous estimates of the effects of ADHD on behavioural and social outcomes and health measures^[21–24]^. However, many of the estimates attenuated to the null (especially for behavioral and social outcomes) when using the family-based Mendelian randomization analysis (**Figure 3**). Results were largely consistent across sensitivity analyses (**eFigure 1**).

**Figure 3:**
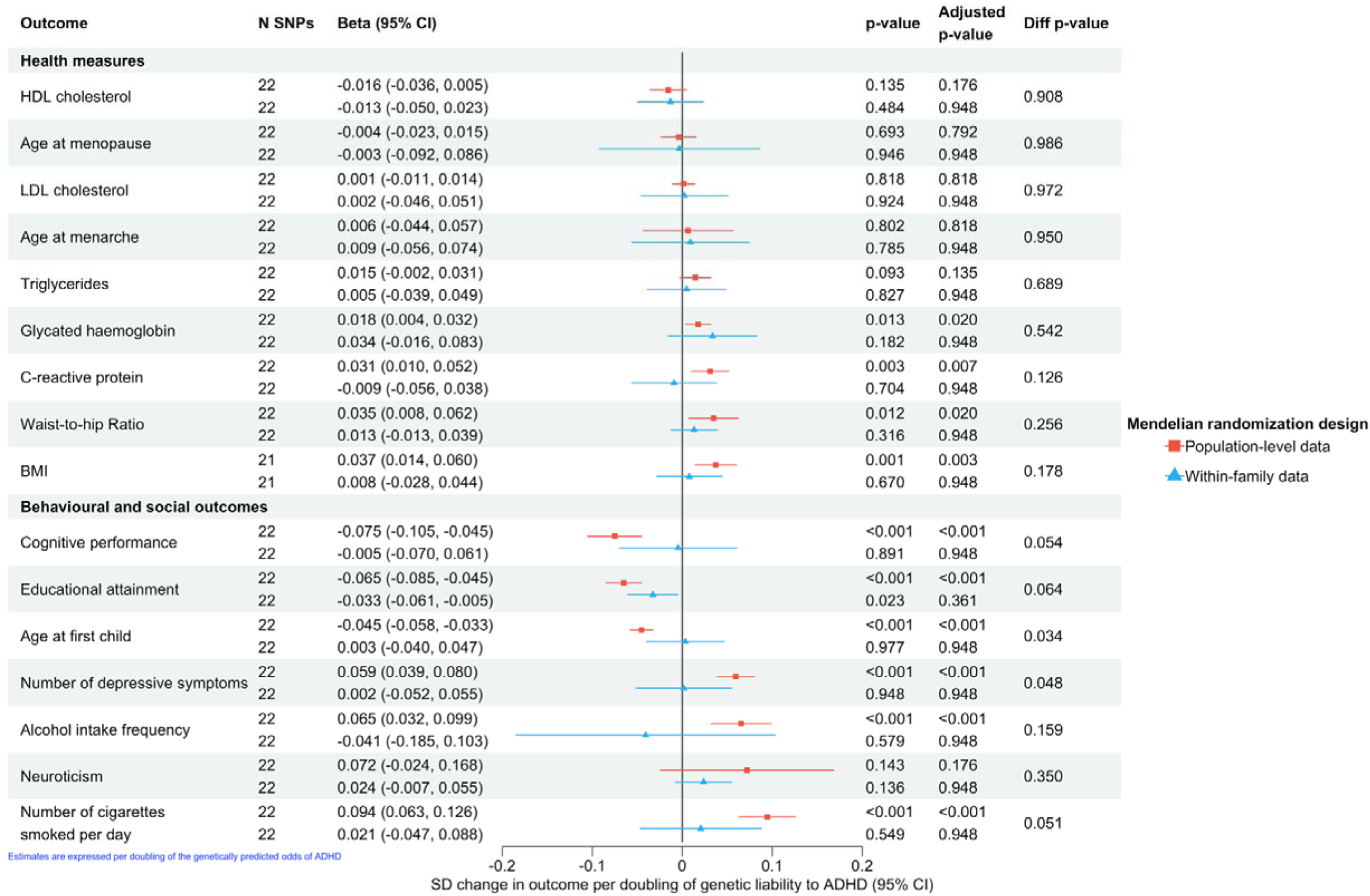
Inverse variance weighted estimates for the association between a genetic liability to ADHD and health outcomes using population-based Mendelian randomization and within-family (sibling pairs) Mendelian randomization. *Nb: poluation-level data refers to a Mendelian randomization analysis conducted using population-level GWAS data, while Within-family data refers to a Mendelian randomization conducted using within-family (sibling-level) data. Adjusted p-value refers to FDR corrected p-value; Diff p-value represents the Wald test for heterogeneity betwwne Mendelian randomization methods*.

### Causal effect of genetic liability to ADHD on behavioural and social outcomes

Population-based Mendelian randomization implied that doubling a person’s genetic liability to ADHD increased: how frequently they drank alcohol by 0.065 SD (95%CI:0.032,0.099); the number of depressive symptoms they experienced by 0.059 SD (95%CI:0.039,0.080); and the number of cigarettes they smoked per day by 0.094 SD (95%CI:0.063,0.126). There was also evidence that doubling a person’s genetic liability to ADHD reduced educational attainment by 0.065 SD (95%CI:-0.085,-0.045), lowered the age at which they had their first child by 0.045 SD (95% CI:-0.058,-0.033), and reduced their cognitive performance by 0.075 SD (95% CI:-0.105,-0.045). These estimators provided little evidence that higher genetic liability to ADHD affected neuroticism after correction for multiple testing.

In contrast, our family-based Mendelian randomization estimates implied that doubling a person’s genetic liability to ADHD had little effect on: how frequently they drank alcohol (beta:-0.041;95%CI:-0.185,0.103), the age at which they had their first child (beta:0.003,95%CI:-0.040,0.047), the number of depressive symptoms they experience (beta:0.002,95% CI:-0.052,0.055), the number of cigarettes they smoke per day (beta:0.021,95%CI:-0.047,0.088) or their cognitive performance (beta:-0.005,95%CI:-0.070,0.061). The effect on educational attainment also attenuated (beta:-0.033,95%CI:-0.061,-0.005).

### Causal effect of a genetic liability to ADHD on health measures

Population-based Mendelian randomization implied that doubling a person’s genetic liability to ADHD increased BMI by 0.037 SD (95%CI: 0.014,0.060), waist-to-hip ratio by 0.035 SD (95%CI:0.008,0.062), glycated haemoglobin levels by 0.018 SD (95%CI:0.004,0.032) and CRP levels by 0.031 SD (95%CI:0.010,0.052). These estimators provided little evidence that higher genetic liability to ADHD affected HDL-cholesterol, LDL-cholesterol, triglycerides, age at menarche, or age at menopause after correction for multiple testing.

In comparison, in the family-based Mendelian randomization estimates provide little evidence that doubling a person’s genetic liability to ADHD affected BMI (beta:0.008, 95%CI:-0.028,0.044), waist-to-hip ratio (beta:0.013, 95%CI:-0.013,0.039), glycated haemoglobin levels (beta: 0.034, 95%CI:-0.016,0.083) or CRP levels (beta: -0.009, 95%CI:-0.056,0.038). The population-based and family-based estimates were statistically consistent for these outcomes, with their confidence intervals overlapping despite differences in point estimates.

Therefore, across the 16 outcomes, population-based estimates reproduced previously reported associations between genetic liability to ADHD and worse health, behavioural and social outcomes (Figure 3). In family-based analyses, these estimates were attenuated for most outcomes and exhibited a trend of differing from the population-based estimates, e.g. for depressive symptoms (p=0·048) and age at first child (p=0·034), cigarettes smoked per day (p=0·051), cognitive performance (p=0·054) and educational attainment (0.064). The remaining outcomes were imprecise and consistent with both the null and the population-based estimate.

Due to constraints around data availability, both the population-based and family-based Mendelian randomization analyses used a SNP-exposure estimate derived from a large population-based GWAS. Therefore, SNP-exposure estimates are subject to confounding from population stratification, dynastic effects, and cross-trait assortative mating. Thus, social and environmental factors may influence the denominator and lead to inconsistent estimates in both designs. To examine this, we re-calculated the IVW estimate after applying a range of shrinkage parameters (0.05-0.4) to the SNP-exposure estimate. These parameters include extreme values to capture both plausible and inflated scenarios. Results were largely unchanged, suggesting that our findings are robust to even substantial inconsistency in the SNP-exposure associations (**eFigure 2**).

In the population-based Mendelian randomization analyses, we observed evidence of heterogeneity across SNPs (Cochran’s Q test) for all phenotypes (except LDL-cholesterol, depressive symptoms, WHR, cigarettes smoked per day and menopause). Heterogeneity was greatest in the neuroticism phenotype (Q=156.27, p=1.12×10^-22^; driven by 5 SNPs: rs11596214, rs4916723, rs549845, rs7506904, and rs77960) which may explain its larger than expected confidence intervals. In the family-based Mendelian randomization, heterogeneity across SNPs was only present for the phenotypes age at menarche (Q=35.37, p=0.048) and BMI (Q=38.42, p =0.037). Results are presented in **eTable 5.**

When evaluating pleiotropy with MR-PRESSO, we detected horizontal pleiotropic variants in a small number of results. However, removal only had a minor effect on the strength of the previously reported relationships. This suggests that while individual SNP-outcome estimates may vary (appear heterogeneous), their differences are unlikely to reflect systematic pleiotropy that would make the overall causal estimate inconsistent. Identified outliers and pleiotropy-corrected associations measured by MR-PRESSO are presented in **eTable 6.** The consistency of results across pleiotropy-robust sensitivity analyses suggests that individual-level SNP-birthweight associations did not drive the Mendelian randomization estimates in either the population-based or family-level Mendelian randomization estimates.

Associations when using the smaller population-based data from the Within Family Consortium were consistent with our main population-based Mendelian randomization analysis. This increases confidence that the observed difference in effect estimates between the population-based and within-family Mendelian randomization designs is due to the adjustment for environmental and social effects and not as a consequence of sample differences (see **eResults**).

## Discussion

We used data from 38,691 people with ADHD, 186,843 controls and up to 164,220 siblings to compare conventional population-based Mendelian randomization estimates of the effects of genetic liability to ADHD on health, behavioural and social outcomes with family-based estimates. Both designs aim to estimate the causal effect of an individual’s genetic liability to ADHD, but population-based estimates are susceptible to confounding by the familial and social environment in which ADHD arises, whereas family-based estimates isolate the effect of an individual’s own liability and are robust to population stratification, assortative mating and dynastic effects (outlined in **Box 1** and **Figure 2**)^[25, 52, 53]^. Population-based estimates reproduced results across the observational and Mendelian randomization literature^[21, 22, 24]^, whereas in family-based analyses, most effects attenuated towards the null, suggesting the population-based estimates for depressive symptoms and age at first child were inconsistent.

These results have important implications for how the harms associated with ADHD are interpreted. The estimated effects of genetic liability to ADHD on depressive symptoms and earlier age at first child, seen in population-based analyses, were not reproduced in family-based analyses, indicating that an individual’s own liability is unlikely to be a major direct cause of these outcomes. Instead, these estimates may reflect the family and social context in which ADHD occurs, most plausibly through confounding of the population-based estimates by population stratification, assortative mating, and/or dynastic effects^[26, 29]^. For the remaining outcomes, the family-based estimates were consistent with both the population-based estimate and the null, and therefore, we cannot determine how much of each association reflects a direct individual-level effect rather than familial or environmental factors.

If part of the behavioural and social burden associated with ADHD reflects the family and environmental context in which ADHD arises, rather than the individual’s own liability, then interventions directed solely at the individual, including pharmacological treatment, may not reduce these outcomes as much as observational and population-based genetic estimates imply. The same distinction has implications for burden-of-disease and health-economic models, which typically attribute these associations causally to ADHD and may overstate the fraction modifiable by treating the individual. Clinical and policy conclusions should be drawn from bodies of evidence, not from single studies^[54]^; future research should rigorously test whether these implications translate into effective interventions.

Our findings extend the existing literature: earlier observational and Mendelian randomization studies could not separate direct from nondirect effects because sufficient family-based data were not available^[26, 27]^. Previously reported population-based estimates reflect real differences in outcomes, but they could not test whether these effects were due to individual-level or broader factors. Our results are consistent with Soler Artigas and colleagues^[22]^, where the two-sample Mendelian randomization analysis highlighted several temporally implausible associations, including genetic liability to smoking apparently increasing the risk of ADHD, which is typically diagnosed years before smoking initiation^[55]^. Such findings can be readily explained by the population-level processes (**Box 1**, **Figure 2**) for which family-based designs can control^[25]^.

Interpretation is complicated by ADHD medication and other treatments. These cannot confound our estimates, as they do not affect germline genetic variation, but they may lie on the causal pathway between ADHD liability and later outcomes. Our estimates, including the family-based estimates, therefore reflect the total effect of genetic liability to ADHD, including any pathways acting through treatment, and cannot isolate the effect of treatment itself.

Our study also has limitations. The SNP–exposure associations were derived from a population-based GWAS and could be influenced by familial factors; however, estimates were essentially unchanged across a wide range of shrinkage parameters, suggesting that inflation of the SNP–ADHD association is unlikely to affect our conclusions. Family-based Mendelian randomization is inherently less precise than population-based analysis, because it uses only within-family variation and genotyped relatives remain scarce^[26, 27]^; this likely explains why we could not detect heterogeneity for some outcomes. Our findings highlight the need for larger family-based GWAS and more family-based non-European samples^[25, 52, 53, 56]^. We also did not use Steiger filtering to assess effect direction, as its validity depends on comparable measurement properties and error across the exposure and outcome. In our setting, the exposure is binary and estimated in a separate sample compared to the outcome, which makes such assumptions unlikely.

In conclusion, the associations between genetic liability to ADHD and behavioural and social outcomes reported in population-based studies appear, at least in part, to reflect the family environments in which ADHD arises rather than the direct effect of an individual’s own liability. These findings suggest that some of the burden attributed to an individual’s ADHD may instead reflect familial and social contexts, and that testing causal claims about the individual-level effects of ADHD liability requires familial data. Population-based Mendelian randomization remains valuable, but family-based designs are among the few approaches that can separate direct effects from familial confounding. Progress will depend on assembling much larger samples of genotyped relatives.

## Supporting information

MR Strobe checklist

## Data Availability

GWAS data are publicly available trhogh the OpenGWAS website (https://gwas.mrcieu.ac.uk). Part of this research was conducted by using data from UKB project ID: 81499 and project: 30418, a major biomedical database, which can be provided by UKB (http://www.ukbiobank.ac.uk/).
Information about the data sources used in this study is summarised in eTable 3. The study relied on summary-level data that have been made publicly available. Ethical approval was obtained for all original studies. Methods of assessment and diagnostic criteria for each exposure/outcome are described in the relevant papers. Full harmonised SNP lists for each Mendelian randomization analysis are available on GitHub

https://github.com/daisycrick/Within_family_ADHD

https://gwas.mrcieu.ac.uk

## Funding

NMD is supported by the Norwegian Research Council: 295989, the Medical Research Council (MRC) MR/V002147/1 and UKRI1510: the National Institute for Mental Health (NIMH) MH130448, and the <u>Division of Psychiatry</u> (https://www.ucl.ac.uk/brain-sciences/psychiatry). SEM is supported by an Australian National Health and Medical Research Council Investigator Grant (2025674). DME and this research is supported by an Australian National Health and Medical Research Council Investigator Grant (2017942).

## Conflict of interest

None declared.

## Acknowledgements

This research has been conducted using data from UK Biobank.

## Ethical Approval

UK Biobank has approval from the North West Multi-centre Research Ethics Committee (MREC) as a Research Tissue Bank (RTB) approval.

## Data sharing, sources and instruments

GWAS data are publicly available trhogh the OpenGWAS website (https://gwas.mrcieu.ac.uk). Part of this research was conducted by using data from UKB project ID: 81499 and project: 30418, a major biomedical database, which can be provided by UKB (http://www.ukbiobank.ac.uk/).

Information about the data sources used in this study is summarised in **eTable 3.** The study relied on summary-level data that have been made publicly available. Ethical approval was obtained for all original studies. Methods of assessment and diagnostic criteria for each exposure/outcome are described in the relevant papers. Full harmonised SNP lists for each Mendelian randomization analysis are available on GitHub (https://github.com/daisycrick/Within_family_ADHD).

## Code availability and software

Code for data management and statistical analysis is available at: https://github.com/daisycrick.

SOFTWARE: R version 4.5.2, Plink1.9; data manipulation: dplyr, tidyverse; Packages: TwoSampleMR, MendelianRandomization, MR-PRESSO; Visualisation: forestplotter, ggplot2.

Study protocol was not pre-registered.

## Reporting Guidelines

This study is reported in accordance with the STROBE-MR (Strengthening the Reporting of Observational Studies in Epidemiology using Mendelian Randomization) guidelines (checklist attached).

## Author Contributions

**Conceptualization:** DCPC, DME, SEM; **Methodology:** DCPC, DME; **Investigation:** DCPC; **Supervision:** DME, ND; **Writing original draft:** DCPC; **Writing - review and editing:** DCPC, DME, ND, SEM.

---

**eFigure 1:**
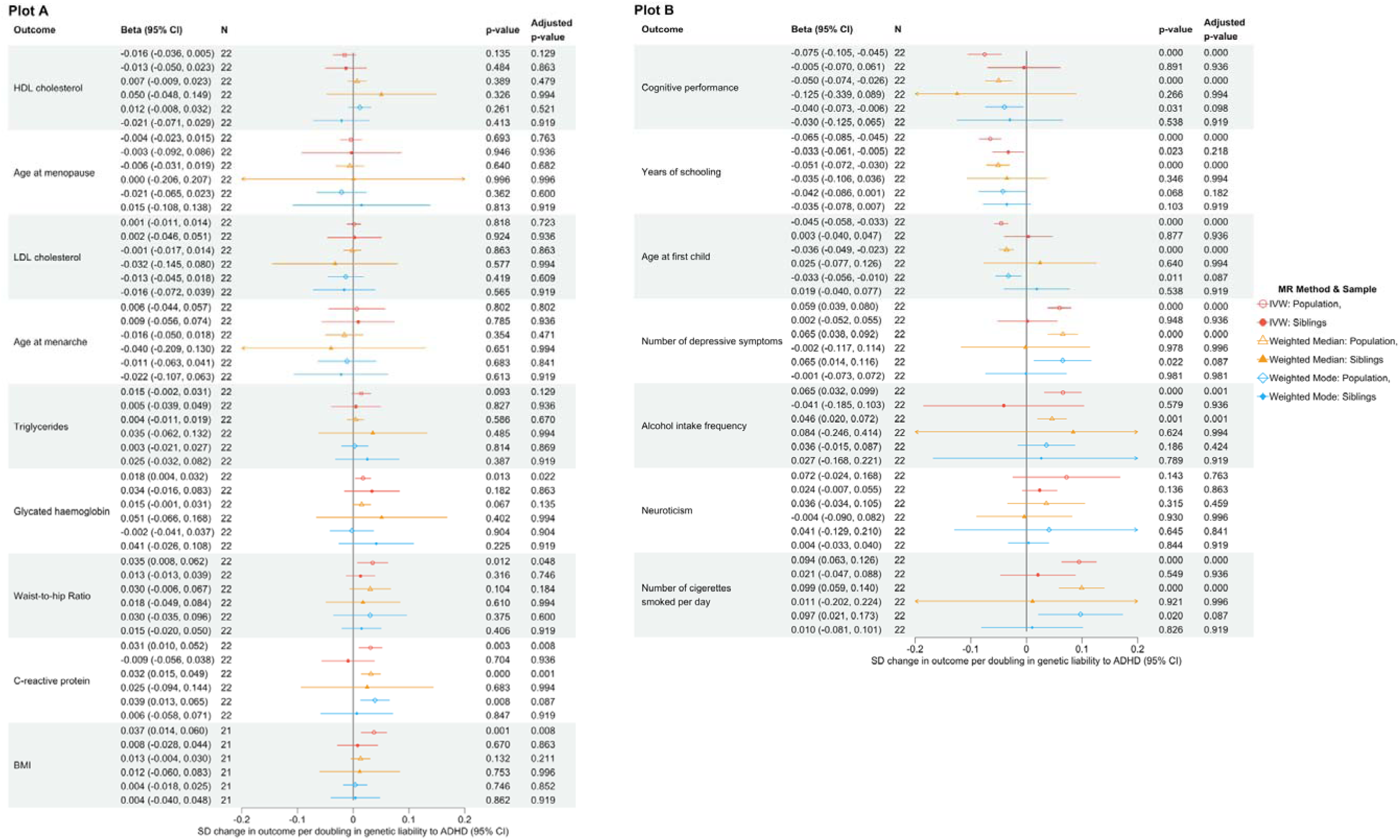
Population-level and Within-family Mendelian randomization (MR) estimates for the effect of genetic liability to ADHD on health and behavioural outcomes using the inverse variance weight method (IVW), the weighted median method and the weight mode method.

**eFigure 2:**
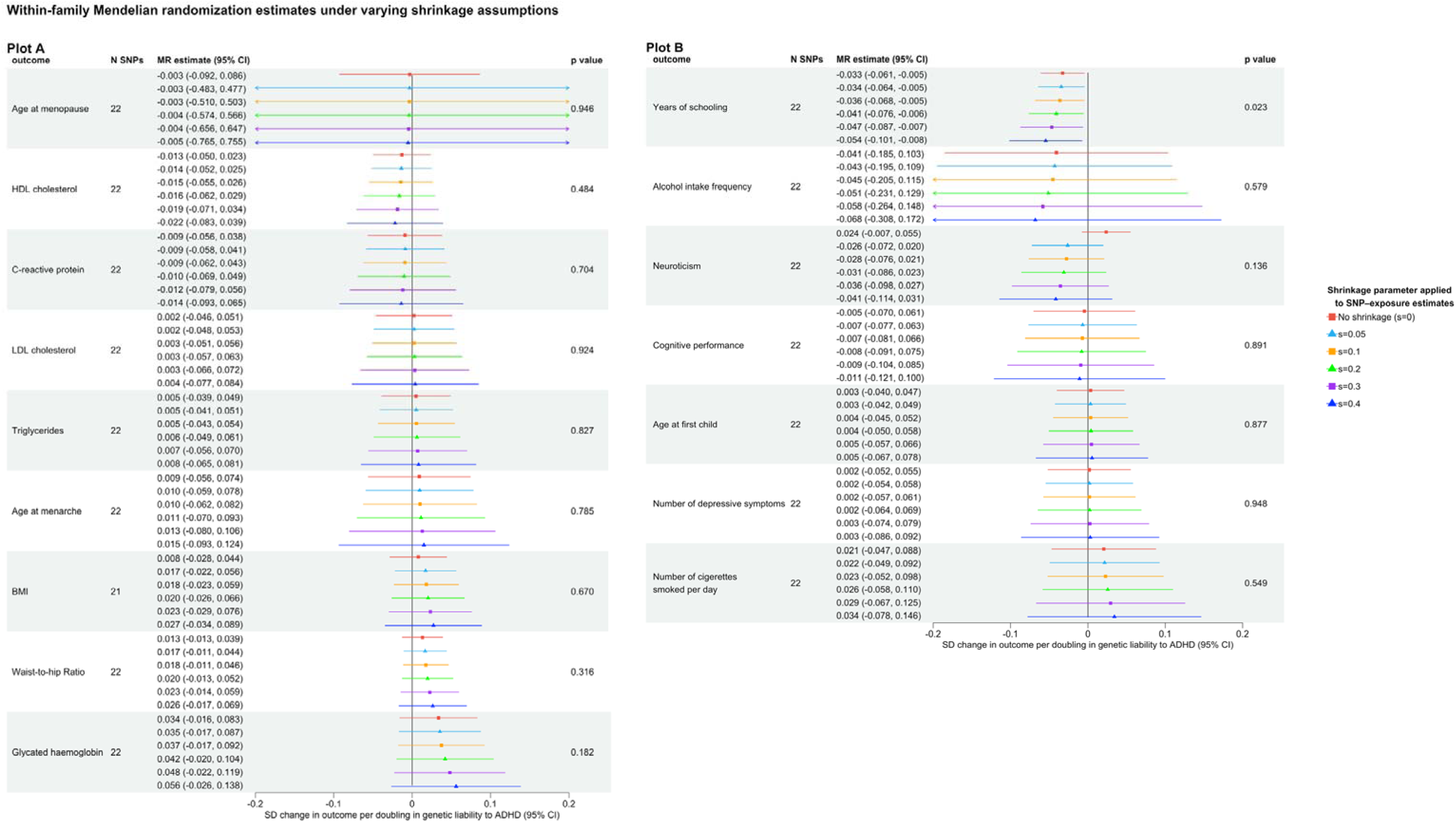
Within-family Mendelian randomization (MR) estimates for the effect of genetic liability to ADHD on biological measures (Plot A) and lifestyle factors (Plot B) under varying levels of shrinkage applied to SNP–exposure associations. *Shrinkage was applied as a method to account for potential inflation in population-based SNP–exposure estimates, which were used as instruments in both the population and within-family MR analyses. SNP–exposure effects were rescaled by **1-s**, where **s** represents the degree of shrinkage*.

## Notes

### Competing Interest Statement

The authors have declared no competing interest.

### Author Declarations

GWAS data are publicly available trhogh the OpenGWAS website (https://gwas.mrcieu.ac.uk).

