## Supplementary tables for "Familial factors inflate the estimated effects of genetic liability to ADHD on behavioural outcomes"

| **eTable 1: Overview of key recent studies investigating the association between a genetic liability to ADHD and health outcomes/behaviours using Mendelian randomization (see list of abbreviations at the foot of the table)** | | | | | |
| --- | --- | --- | --- | --- | --- |
| Study | Year | Main causal associations found | Main MR method | Notes | Address confounding/violation of the Independence assumption |
| Mediating pathways between attention deficit hyperactivity disorder and type 2 diabetes mellitus: evidence from a two-step and multivariable Mendelian randomization study^[1]^ | 2024 | + T2D | Two-step two-sample Mendelian randomization and MVMR | Three of nine purported mediators were found to have mediation effects: educational attainment, BMI and television watching. | No |
| Bidirectional associations between mental disorders, antidepressants and cardiovascular disease^[2]^ | 2024 | + CAD, CVD and heart failure | Two-sample MR and MVMR | ADHD strongly increases the risk of CVD, but CVD has little causal effect on mental disorders. | No |
| Understanding the causal relationships of attention deficit/hyperactivity disorder with mental disorders and suicide attempt: a network Mendelian randomization study^[3]^ | 2023 | + Anorexia nervosa, MDD, suicide attempt and PTSD | Two-sample network Mendelian randomization analysis and MVMR | Provides evidence about directionality rather than effect size due to the use of a smaller set of ADHD instruments (n=12). | No |
| Shared genetics and bidirectional causal relationships between type 2 diabetes and attention-deficit/hyperactivity disorder^[4]^ | 2023 | + T2D | Two-sample MR |  | No |
| Mendelian randomization analysis for attention deficit/hyperactivity disorder: studying a broad range of exposures and outcomes^[5]^ | 2023 | + MDD, smoking behaviour, number of sexual partners, time spent watching television.  - age at first sexual intercourse, age completed full-time education and years of schooling. | Two-sample MR | Investigated 124 lifestyle/health-relevant traits. | No |
| Understanding the causal relationships of attention-deficit/hyperactivity disorder with mental disorders and suicide attempt: a network Mendelian randomisation study^[6]^ | 2023 | + AN, MDD, Suicide attempt and PTSD | Two-sample network MR | Direct association between a genetic liability for ADHD and suicide attempts and ADHD and PTSD after adjustment for MDD. | No |
| The Relationship of Attention-Deficit/Hyperactivity Disorder With Posttraumatic Stress Disorder: A Two-Sample Mendelian Randomization and Population-Based Sibling Comparison Study^[7]^ | 2023 | + PTSD | Two-sample MR and a population-based sibling comparison (Cox model with separate strata for each cluster of siblings) | Although the study found that there was an increased risk for developing PTSD in individuals with ADHD compared to undiagnosed siblings. The sibling comparison design did not investigate if this relationship was causal. | Yes (although not when investigating causality) |
| Attention-deficit/hyperactivity disorder and ischemic stroke: A Mendelian randomization study^[8]^ | 2023 | + Ischemic stroke, large-artery atherosclerotic stroke | Two-sample MR | Associations persisted in sensitivity analyses and MVMR. Sparse ADHD instruments (n=13). | No |
| The Effect of Attention Deficit/Hyperactivity Disorder on Physical Health Outcomes: A 2-Sample Mendelian Randomization Study^[9]^ | 2021 | + Childhood obesity, CAD | Two-sample MR | Health outcomes included metabolic, cardiovascular and autoimmune endpoints. The study used older ADHD instruments with limited precision (n=13). Most of the physical outcomes investigated remain null/inconclusive.  The relationship between ADHD and CAD attenuated when controlling for childhood obesity. | No |
| ADHD and depression: investigating a causal explanation^[10]^ | 2021 | + Recurrent depression in later life, and MDD. | Two-sample MR | Broader depression phenotypes behaved differently so the finding is only applicable to the narrowly defined MDD. | No |
| The link between attention deficit hyperactivity disorder (ADHD) symptoms and obesity-related traits: genetic and prenatal explanations^[11]^ | 2021 | + Obesity-related traits (BMI, WC, WHR (unadjusted and adjusted for BMI) body fat % and BMR). | Two-sample MR |  | No |
| Investigating causality between liability to ADHD and substance use, and liability to substance use and ADHD risk, using Mendelian randomization^[12]^ | 2019 | + Smoking initiation, heaviness of smoking among smokers and cannabis initiation.  - likelihood of smoking cessation. | Two-sample MR |  | No |
| **Studies identified in the literature search investigating ADHD, Mendelian randomization, and within-family study designs** | | | | | |
| Study | Year | Main causal associations found | Methods/designs | Notes | Address confounding/violation of the Independence assumption |
| Gene-environment correlations and causal effects of childhood maltreatment on physical and mental health: a genetically informed approach^[13]^ | 2021 | Evidence consistent with a causal relationship between childhood maltreatment and depression, and bidirectional relationships with schizophrenia and ADHD.  No causal role of childhood maltreatment on coronary artery disease, type 2 diabetes or CRP levels. | GWAS meta-analysis, single nucleotide polymorphism heritability, genetic correlations, family-based and population-based polygenic scores | Family-based PGS analyses and population-based MR were conducted separately. | No |
| Body mass index and childhood symptoms of depression, anxiety and attention-deficit hyperactivity disorder: A within-family Mendelian randomization study^[14]^ | 2022 | Own BMI increase depressive symptoms and ADHD symptoms. Maternal BMI may independently affect child’s depressive symptoms.  Within family MR did not support an impact of BMI on these outcomes. | One sample trio MR and within-family MR | Within-family MR controlling for parental genotype.  Results differed according to the genetic instrument and did not provide consistent evidence. | Yes |
| Testing maternal effect of vitamin-D and omega-3 levels on offspring neurodevelopmental traits in the Norwegian Mother, father and child cohort study^[15]^ | 2024 | Effects of maternal vitamin D levels on ADHD traits did not remain after controlling for familial genetic predisposition using trio-MR. | Two-sample MR and mother-father-child trio MR | Mother-father-child trio MR, including paternal negative control. | Yes |
| The relationship of Attention-deficit/Hyperactivity Disorder with posttraumatic Stress disorder: A two sample Mendelian randomization and population-based sibling comparison study^[7]^ | 2023 | ADHD genetic liability causally linked to increased risk for PTSD. Individuals diagnosed with ADHD were at higher risk for developing PTSD than their undiagnosed sibling. | Two-sample MR. | Population-based with sibling comparison. The family design supported the ADHD-> PTSD association. | No |
| Attention-Deficit/Hyperactivity Disorder and Major Depressive Disorder: Evidence from multiple genetically informed designs^[16]^ | 2024 | MR analyses showed that genetic liability for ADHD is causally related to MDD. Sibling and twin comparisons indicated that individuals with ADHD have increased risk for subsequent development of MDD. | Generalised linear model, cox proportional hazards models adjusted for genetic and environmental factors shared by siblings. Two-sample MR | Population-based MR plus separate sibling and co-twin comparison. | No |
| Parental education and children’s depression, anxiety and ADHD traits, a within-family study in MoBa^[17]^ | Preprint | Little evidence that mothers’ or father’s educational attainment independently affected children’s depressive, anxiety, or ADHD traits. | One sample trio MR and supplementary two-sample MR | Mother-father-offspring trios using MoBa and therefore small samples. | Yes |
| Are there causal relationships between attention-deficit/hyperactivity disorder and body mass index? Evidence from multiple genetically informed designs^[18]^ | 2021 | Bidirectional relationship between ADHD and BMI. MZ twin-difference analysis provided little support of this. | Two-sample bidirectional MR.  MZ cross-lagged twin difference3s and within-/between-family polygenic score. | Population-based MR plus twin-difference and within-family PGS analyses. | No |
| Evaluating the causal relationship between educational attainment and mental health^[19]^ | 2024 | Lower educational attainment increases risk of MDD, ADHD, alcohol dependence, GAD and PTSD. There was also a causal effect of ADHD on lower educational attainment. | Two sample MR | Population-based MR plus a separate within-sibship analysis. | No |
| NB: Table features a selection of the key recent studies that investigate the causal relationship between ADHD and health outcomes. It is not exhaustible but demonstrates how little has been done to control for population stratification, assortative mating and dynastic effects.  T2D= Type 2 diabetes, BMI = body mass index, CAD= coronary artery disease, CVD = cardiovascular disease, MR= Mendelian randomization, MVMR = Multivariable Mendelian randomization, MDD = major depressive disorder, PTSD = post-traumatic stress disorder, ADHD = attention, deficit hyperactivity disorder, AN= anorexia Nervosa, WC = waist circumference, WHR = waist-to-hip ratio, BMR = basal metabolic rate, GAD = generalised anxiety disorder. | | | | | |

| **eTable 2: Genome-wide significant SNPs used as instrumental variables, their closest gene and putative function** | | | | | | | | | | |
| --- | --- | --- | --- | --- | --- | --- | --- | --- | --- | --- |
| Chr | Pos | RSID | EA | OA | EAF | Beta (SE) | P | Closest Gene | Putative gene Function | Known association |
| 1 | 44076469 | rs549845 | G | A | 0.321 | 0.079 (0.01) | 9.03E-15 | *PTPRF* | Possible cell adhesion receptor. The gene encodes an enzyme that acts as a receptor-linked protein tyrosine phosphatase. Regulates a variety of cellular processes including cell growth, differentiation, mitotic cycle and oncogenic transformation. Also shown to function in the regulation of epithelial cell-cell contacts at adherents junctions and in the control of beta-catenin signalling. | ADHD, intelligence, smoking behaviour, educational attainment, mental or behavioural disorder |
| 2 | 145714354 | rs1438898 | A | C | 0.762 | 0.063 (0.01) | 4.88E-09 | *TEX41* | An RNA gene affiliated with the IncRNA class. | Risk-taking behaviour, aging rate, haemoglobin measurement |
| 3 | 51884072 | rs2311059 | G | A | 0.314 | -0.058 (0.01) | 3.16E-08 | *IQCF2, IQCF3* | ***IQCF2*** and ***IQCF3***  are protein coding genes predicted to enable calmodulin binding activity.. | ADHD, risk-taking behaviour |
| 3 | 20724204 | rs2886697 | G | A | 0.634 | 0.059 (0.01) | 7.90E-10 | *SGO1-AS1* | An RNA gene affiliated with the IncRNA class. | ADHD, risk-taking behaviour |
| 3 | 71499401 | rs17718444 | C | T | 0.695 | 0.061 (0.01) | 2.87E-09 | *FOXP1, EIF4E3, MIR1284, FOXP1-IT1, ENSG00000285708, Inc-GPR27-16, HSALNG0026758* | ***FOXP1*** plays important roles in the regulation of tissue and cell type-specific gene transcription during both development and adulthood. May act as a tumour suppressor. It plays an important role in the specification and differentiation of lung epithelium. ***EIF4E3*** is a protein coding gene, and can recognise and binds the 7-methylguanosine mRNA cap during an early step in the initiation of protein synthesis. ***MIR1284*** is affiliated with the miRNA class. ***FOXP1-IT1*** is affiliated with the IncRNA class. **ENSG00000285708** (novel protein) is a protein coding gene. ***Inc-GPR27-16*** is an RNA gene affiliated with the IncRNA class. ***HSALNG0026758*** is an RNA Gene affiliated with the IncRNA class. | Risk-taking behaviour, nasal cavity polyp, ADHD, level of zona pellucida sperm-binding protein 3 in blood |
| 4 | 147099654 | rs6537401 | G | A | 0.66 | -0.057 (0.01) | 1.40E-08 | *LSM6* | LSM6 plays a role in pre-mRNA splicing as component of the U4/U6-U5 tri-snRNP complex that is involved in the spliceosome assembly. | ADHD, risk-taking behaviour |
| 4 | 112217523 | rs17576773 | C | T | 0.888 | 0.096 (0.02) | 1.63E-10 | *RNU-289P, LYPLA1P2* | Pseudogenes | ADHD, risk-taking behaviour |
| 5 | 144474779 | rs10875612 | C | T | 0.483 | -0.054 (0.01) | 5.62E-09 | *ASS1P10, NAMPTP2* | Pseudogenes | ADHD, risk-taking behaviour |
| 5 | 87854395 | rs4916723 | A | C | 0.553 | -0.085 (0.01) | 9.48E-15 | *MIR9-2HG, MIR34C, MIR9-2* | ***MIR9-2HG*** is an RNA gene affiliated with the IncRNA class. It is an evolutionary conserved gene that produces alternatively spliced long non-coding RNAs that may be expressed predominantly in the brain and visual cortes. Transcripts may be involved in tumorigenesis, as depletion by siRNA suppressed glioma cell division. ***MIR34C*** is an RNA gene and is affiliated with the miRNA class. ***MIR9-2*** is an RNA gene and is affiliated with the miRNA class. | ADHD, alcohol consumption quality, ASD, MDD, bipolar disorder |
| 5 | 103964585 | rs77960 | G | A | 0.665 | -0.073 (0.01) | 2.46E-13 | *NIHCOLE, RNU6-334P* | ***NIHCOLE*** is an RNA gene and is affiliated with the IncRNA class. ***RNU6*-*334P*** is a pseudogene. | MDD, stress-related disorder, anxiety, risk-taking behaviour, chronotype measurement, mathematical ability |
| 6 | 70858701 | rs2025286 | A | C | 0.553 | -0.055 (0.01) | 4.00E-09 | *COL19A1* | May act as a cross-bridge between fibrils and other extracellular matrix molecules. Involved in skeletal myogenesis in the developing oesophagus and may play a role in organization of the pericellular matrix of the sphincteric smooth muscle. | Risk taking, ADHD |
| 7 | 114158954 | rs9969232 | G | A | 0.344 | -0.068 (0.01) | 9.98E-12 | *FOXP2* | Transcriptional repressor that may play a role in the specification and differentiation of lung epithelium. May also play a role in developing neural, gastrointestinal and cardiovascular tissues. The protein contains FOX DNA-binding domain and a large polyglutamine tract and is evolutionarily conserved transcription factor which may bind to ~ 300-400 gene promoters in the human genome to regulate the expression of a variety of genes. It is required for the proper development of speech and language regions of the brain during embryogenesis and may be involved in a variety of biological pathways and cascades that may ultimately influence language development. | Household income, ADHD |
| 7 | 67685754 | rs73145587 | A | T | 0.91 | 0.101(0.02) | 3.67E-08 | *LOC105375341, MTND4P3, MTCO1P57* | ***LOC105375341*** is an RNA gene and is affiliated with the IncRNA class. ***MTND4P3*** is a pseudogene. ***MTCO1P57*** is a pseudogene. | ADHD, risk-taking behaviour |
| 8 | 93277087 | rs7844069 | T | G | 0.428 | 0.055 (0.01) | 6.74E-09 | *RUNX1T1, FLJ46284* | ***RUNX1T1*** encodes a member of the myeloid translocation gene family which interact with DNA-bound transcription factors and recruit a range of corepressors to facilitate transcriptional repression. Among its related pathways are wnt/beta-catenin signalling pathway in leukaemia and acute myeloid leukaemia. ***FLJ46284*** is an RNA gene and is affiliated with the IncRNA class. | Insomnia, risk-taking behaviour, ADHD |
| 8 | 145802447 | rs4925811 | T | G | 0.515 | -0.058 (0.01) | 8.30E-09 | *ARHGAP39* | Predicted to enable GTPase activator activity. It is involved in post synapse organization and is active in glutamatergic synapse. Among its related pathways are nervous system development and signalling by Rho GTPases. | ADHD, risk-taking behaviour |
| 10 | 106453832 | rs11596214 | G | A | 0.597 | 0.053 (0.01) | 3.17E-08 | *SORCS3* | This gene encodes a type-1 receptor transmembrane protein that is a member of the vacuolar protein sorting 10 receptor family. Genetic variation in this gene is associated with Alzheimer’s disease. Knockdown of the gene in cell culture results in an increase in amyloid precursor protein processing. | ADHD, MDD, neuroticism measurement, social inhibition quality, substance abuse |
| 10 | 8784773 | rs11255890 | C | A | 0.389 | 0.053 (0.01) | 4.14E-08 | *LINC02676, RNA5SP299* | ***LINC02676*** is an RNA gene is affiliated with the IncRNA class. ***RNA5SP299*** is a pseudogene. |  |
| 11 | 28602173 | rs2582895 | C | A | 0.634 | 0.072 (0.01) | 4.09E-14 | *LINC02758* | RNA gene affiliated with the IncRNA class. |  |
| 12 | 89771903 | rs704061 | T | C | 0.554 | -0.056 (0.01) | 2.30E-09 |  |  |  |
| 14 | 98690923 | rs76284431 | T | A | 0.847 | -0.082 (0.01) | 1.19E-09 | *LOC105370656, LINC02295, RN7SL714P* | ***LOC105370656*** is an RNA gene affiliated with the ncRNA class. ***LINC02295*** is an RNA gene affiliated with the IncRNA class. ***RN7SL714P*** is a pseudogene. | Risk-taking behaviour, sexual ability behaviour attribute |
| 16 | 61966703 | rs1162202 | C | T | 0.63 | 0.061 (0.01) | 1.92E-09 | *CDH8-AS1, CDH8* | ***CDH8-AS1*** is an RNA gene affiliated with the IncRNA class. ***CDH8*** encodes a type 2 classical cadherin from the cadherin superfamily, integral membrane proteins that mediate calcium-dependent cell-cell adhesion. This cadherin is expressed in brain and is putatively involved in synaptic adhesion, axon outgrowth and guidance. | ADHD |
| 18 | 5871800 | rs76857496 | C | A | 0.87 | 0.080 (0.01) | 1.24E-08 | *MIR3976HG* | RNA gene affiliated with the IncRNA class. | Risk-taking, ADHD |
| 18 | 50625779 | rs7506904 | G | A | 0.343 | -0.056 (0.01) | 1.24E-08 | *DCC* | This gene encodes a netrin 1 receptor. The transmembrane protein is a member of the immunoglobin superfamily of cell adhesion molecules and mediates axon guidance of neuronal growth condes towards sources of netrin 1 ligand. It mediates axon attraction of neuronal growth cones in the developing neurons system upon ligand binding. Its association with UNC5 proteins may trigger signalling for axon repulsion. It also acts as a dependence receptor requires for apoptosis induction when not associated with netrin ligand. It has been implicated as a tumour suppressor gene. |  |
| 20 | 21250843 | rs6082363 | T | C | 0.296 | 0.070 (0.01) | 4.38E-12 | *XRN2, ZNF877P, HSALNG0129101* | ***XRN2*** encodes a 5’-3’ exonuclease that promotes transcription termination at cotransciptional cleavage sites. Alternative splicing results in multiple transcript variants encoding different isoforms. It may promote the termination of transcription by RNA polymerase 2. Among its related pathways are rRNA processing in the nucleus and cytosol and Chaperonin-mediated protein folding. ***ZNF877P*** is a pseudogene. ***HSALNG0129101*** is an RNA gene and is affiliated with the IncRNA class. | ADHD, risk-taking behaviour |
| *NB: Information of Putative Gene function come from websites UniProt, GeneCards, RefSeq and NCBI Gene. Genes with no putative function had no available information.  Beta refers to the effect allele.* | | | | | | | | | | |

| **eTable 3: Overview of data sources and sample sizes used in the study including raw units and population SD used in conversions.** | | | | | | |
| --- | --- | --- | --- | --- | --- | --- |
| **Phenotype** | **Sample** | **N** | **Consortium** | **IEU code** | **Units** | **Population SD** |
| Body mass index | Population | 681,275 | GIANT and UKB^[20]^ | ieu-b-40 | SD | 4.79 |
|  | Sibling pairs | 51,396 | Within-family Consortium | ieu-b-4815 | kg/m2 |  |
| Waist-Hip Ratio | Population | 124,591 | GIANT^[21]^ | ieu-a-74 | SD | 0.08 |
|  | Sibling pairs | 31,740 | Within-family Consortium | ieu-b-4829 | ratio |  |
| HDL-cholesterol | Population | 403,943 | UK Biobank^[22]^ | ieu-b-109 | SD |  |
|  | Sibling pairs | 37,120 | Within-family Consortium | ieu-b-4843 | SD |  |
| LDL-cholesterol | Population | 440,546 | UK Biobank^[22]^ | ieu-b-110 | SD |  |
|  | Sibling pairs | 29,121 | Within-family Consortium | ieu-b-4845 | SD |  |
| Triglycerides | Population | 441,016 | UK Biobank^[22]^ | ieu-b-111 | SD |  |
|  | Sibling pairs | 30,515 | Within-family Consortium | ieu-b-4849 | SD |  |
| C-reactive protein | Population | 575,531 | UKB + CHARGE^[23]^ | ebi-a-GCST90029070 | SD |  |
|  | Sibling pairs | 20,623 | Within-family Consortium | ieu-b-4763 | SD |  |
| Glycated haemoglobin | Population | 389,889 | UKB^[24]^ | ebi-a-GCST90014006 | SD |  |
|  | Sibling pairs | 17,724 | Within-family Consortium | ieu-b-4841 | SD |  |
| Alcohol consumption | Population | 462,246 | MRC-IEU using PHESANT^[25]^ | ukb-b-5779 | SD |  |
|  | Sibling pairs | 29,540 | Within-family Consortium | ieu-b-4833 | SD |  |
| Cigarettes smoked per day | Population | 249,752 | GSCAN^[26]^ | ieu-b-142 | SD | 9.62 |
|  | Sibling pairs | 9000 | Within-family Consortium | ieu-b-4825 | Cigarettes per day |  |
| Years of schooling | Population | 293,723 | SSGAC^[27]^ | ieu-a-1001 | SD |  |
|  | Sibling pairs | 54,986 | Within-family Consortium | ieu-b-4835 | SD |  |
| Age at first birth | Population | 542,901 | Meta-analysis of 36 studies ^[28]^ | ebi-a-GCST90000050 | Years | 5.91 |
|  | Sibling pairs | 9,999 | Within-family Consortium | ieu-b-4819 | Years |  |
| Age at menarche | Population | 252,514 | ReproGen studies, UKB and 23andMe^[29]^ | ieu-b-5106 | SD | 1.62 |
|  | Sibling pairs | 29,346 | Within-family Consortium | ieu-b-4821 | Years |  |
| Age at menopause | Population | 143,819 | MRC-IEU using PHESANT^[25]^ | ukb-b-17422 | SD | 5.10 |
|  | Sibling pairs | 4,476 | Within-family Consortium | ieu-b-4823 | Years |  |
| Neuroticism | Population | 374,323 | UKB ^[30]^ | ebi-a-GCST90029028 | SD |  |
|  | Sibling pairs | 28,130 | Within-family Consortium | ieu-b-4848 | SD |  |
| Depressive symptoms | Population | 161, 460 | SSGAC^[27]^ | ieu-a-1000 | SD |  |
|  | Sibling pairs | 16,782 | Within-family Consortium | ieu-b-4839 | SD |  |
| Cognitive function | Population | 269,867 | Meta analysis of 14 cohorts^[31]^ | ebi-a-GCST006250 | SD |  |
|  | Sibling pairs | 9,997 | Within-family Consortium | ieu-b-4837 | SD |  |
| *NB:* GWAS were restricted to European genetic ancestry. All variables other than menopause and menarche were conducted in sex-combined GWAS. Menopause and menarche GWAS were conducted in females only.  Population SD is presented for only the phenotypes that underwent transformation. Transformation was needed to convert effect estimates to standard deviation (SD) units. To achieve this, the Mendelian randomization estimates (β and SE) originally expressed in raw units were divided by the population standard deviation of the phenotype. This was to aid with interpretation and comparability across the Mendelian randomization designs | | | | | | |

| **eTable 4: Association between IVs for ADHD and potential confounders of the ADHD-health outcome association** | | | | | | | | |
| --- | --- | --- | --- | --- | --- | --- | --- | --- |
|  | **ADHD** | | **Maternal smoking during pregnancy** | | | **Own Birthweight** | | |
| Exposure | Effect allele | logOR | logOR | 95% CI | *p-value* | beta | 95% CI | *p-value* |
| rs549845 | G | 0.079 | 0.0002 (0.003) | -0.006, 0.006 | 0.948 | 0.027 (0.007) | 0.013, 0.041 | **<0.001** |
| rs1438898 | A | 0.063 | 0.010 (0.003) | 0.004, 0.016 | **0.0004** | 0.013 (0.007) | -0.001, 0.027 | 0.083 |
| rs2886697 | G | 0.059 | -0.002 (0.002) | -0.006, 0.002 | 0.443 | -0.005 (0.007) | -0.019, 0.009 | 0.490 |
| rs2311059 | A | 0.058 | -0.002 (0.003) | -0.008, 0.003 | 0.395 | -0.021 (0.007) | -0.035, 0.007 | **0.002** |
| rs17718444 | C | 0.061 | -0.005 (0.002) | -0.009, -0.001 | 0.037 | 0.006 (0.007) | -0.008, 0.020 | 0.420 |
| rs17576773 | C | 0.096 | -0.004 (0.003) | -0.010, 0.002 | 0.299 | 0.017 (0.010) | -0.003, 0.037 | 0.076 |
| rs6537401 | A | 0.057 | 0.001 (0.002) | -0.003, 0.005 | 0.747 | -0.018 (0.007) | -0.032, -0.004 | **0.009** |
| rs4916723 | C | 0.085 | 0.006 (0.002) | 0.002, 0.010 | 0.019 | -0.011 (0.006) | -0.023, 0.001 | 0.102 |
| rs77960 | A | 0.073 | 0.001 (0.002) | -0.003, 0.005 | 0.819 | -0.007 (0.007) | -0.021, 0.007 | 0.275 |
| rs10875612 | T | 0.054 | 0.001 (0.002) | -0.003, 0.005 | 0.539 | -0.005 (0.006) | -0.017, 0.007 | 0.399 |
| rs2025286 | C | 0.055 | -0.0001 (0.002) | -0.004, 0.004 | 0.964 | -0.006 (0.006) | -0.018, 0.006 | 0.387 |
| rs9969232 | A | 0.068 | 0.005 (0.002) | 0.001, 0.009 | 0.050 | -0.026 (0.007) | -0.040, -0.01 | **<0.001** |
| rs7844069 | T | 0.055 | -0.0002 (0.002) | -0.004, 0.004 | 0.920 | 0.004 (0.006) | -0.008, 0.016 | 0.497 |
| rs4925811 | G | 0.057 | 0.002 (0.002) | -0.002, 0.006 | 0.463 | -0.003 (0.006) | -0.015, 0.009 | 0.593 |
| rs11255890 | C | 0.053 | 0.001 (0.002) | -0.003, 0.005 | 0.815 | -0.001 (0.007) | -0.015, 0.013 | 0.853 |
| rs11596214 | G | 0.053 | -0.001 (0.002) | -0.005, 0.003 | 0.713 | 0.021 (0.007) | 0.007, 0.035 | **0.001** |
| rs2582895 | C | 0.072 | -0.001 (0.002) | -0.005, 0.003 | 0.689 | 0.027 (0.007) | 0.013, 0.041 | **<0.001** |
| rs704061 | C | 0.056 | 0.003 (0.002) | -0.001, 0.007 | 0.259 | -0.015 (0.006) | -0.027, -0.003 | 0.019 |
| rs1162202 | C | 0.061 | 0.0003 (0.002) | -0.004, 0.004 | 0.905 | -0.009 (0.007) | -0.023, 0.005 | 0.188 |
| rs76857496 | C | 0.080 | 0.003 (0.003) | -0.003, 0.009 | 0.424 | 0.005 (0.009) | -0.013, 0.023 | 0.562 |
| rs7506904 | A | 0.056 | -0.001 (0.002) | -0.005, 0.003 | 0.544 | -0.005 (0.007) | -0.019, 0.009 | 0.430 |
| rs6082363 | T | 0.070 | 0.005 (0.003) | -0.001, 0.011 | 0.034 | 0.013 (0.007) | -0.001, 0.027 | 0.062 |
| NB: p-values in bold remain significant after Bonferroni correction | | | | | | | | |

| **eTable 5: Cochran Q results for heterogeneity in the population-based Mendelian randomization using unrelated individuals and the within-family Mendelian randomization using sibling pairs** | | | | | | |
| --- | --- | --- | --- | --- | --- | --- |
|  | Population-based Mendelian randomization using unrelated individuals | | Within-family Mendelian randomization in sibling pairs | | Within Family Cohort Population-level Mendelian randomization | |
|  | Q | p-value | Q | p-value | Q | p-value |
| **Biological measures** | | | | |  |  |
| BMI | 147.36 | 2.01x10^-21^ | 30.82 | 0.058 | 56.77 | <0.001 |
| Waist-to-hip ratio | 26.08 | 0.203 | 16.60 | 0.725 | 26.22 | 0.198 |
| HDL cholesterol | 103.11 | 8.09x10^-13^ | 25.03 | 0.246 | 33.89 | 0.037 |
| LDL cholesterol | 31.66 | 0.063 | 33.83 | 0.038 | 17.94 | 0.653 |
| Triglycerides | 65.36 | 1.91x10^-6^ | 27.24 | 0.163 | 32.41 | 0.053 |
| HbA1c | 42.82 | 0.003 | 21.37 | 0.436 | 32.01 | 0.058 |
| CRP | 97.51 | 7.94x10^-12^ | 14.96 | 0.824 | 23.12 | 0.338 |
| Age at menarche | 127.64 | 2.78x10^-17^ | 26.66 | 0.182 | 31.70 | 0.063 |
| Age at menopause | 24.84 | 0.254 | 21.74 | 0.415 | 27.14 | 0.166 |
| **Lifestyle factors** | | | | |  |  |
| Cigarettes smoked per day | 28.82 | 0.118 | 25.07 | 0.244 | 13.03 | 0.908 |
| Alcohol consumption | 109.48 | 5.85x10^-15^ | 23.39 | 0.324 | 35.47 | 0.025 |
| Years of schooling | 55.74 | 5.51x10^-6^ | 21.12 | 0.452 | 21.49 | 0.429 |
| Age had first child | 59.23 | 1.67x10^-5^ | 12.65 | 0.900 | 24.83 | 0.255 |
| Depressive symptoms | 28.27 | 0.133 | 19.14 | 0.576 | 29.18 | 0.110 |
| Intelligence | 104.06 | 5.47x10^-13^ | 23.56 | 0.315 | 29.82 | 0.096 |
| Neuroticism | 156.27 | 1.12x10^-22^ | 34.92 | 0.029 | 32.30 | 0.055 |

| **eTable 6: MR-PRESSO results for the effect of ADHD on related phenotypes using population and within-family Mendelian randomization using sibling pairs.** | | | | | | |
| --- | --- | --- | --- | --- | --- | --- |
|  | Mendelian randomization  design | Number of Outliers | Global test | Distortion Test | IVW results after outlier correction | |
|  | |  | p-value | p-value | IVW beta (95% CI) | p-value |
| **Biological measures** | | | |  |  |  |
| BMI | Population-based | 4 | <0.001 | 0.474 | 0.064 (0.036, 0.093) | 0.004 |
| HDL | Population-based | 3 | <0.001 | 0.027 | -0.005 (-0.027, 0.017) | 0.651 |
|  | WFC Population-level | 1 | 0.035 | 0.174 | -0.008 (-0.046, 0.030) | 0.676 |
| Triglycerides | Population-based | 2 | <0.001 | 0.6995 | 0.016 (-0.002, 0.034) | 0.095 |
| CRP | Population-based | 3 | <0.001 | 0.231 | 0.066 (0.046, 0.087) | 5.07x10^-6^ |
| Menarche | Population-based | 3 | <0.001 | 0.249 | -0.018 (-0.061, 0.025) | 0.429 |
| **Lifestyle factors** | |  |  |  |  |  |
| Alcohol consumption | Population-based | 5 | <0.001 | 0.0366 | 0.064 (0.033, 0.096) | 0.001 |
|  | WFC population-level | 1 | 0.028 | 0.719 | 0.073 (-0.066, 0.212) | 0.317 |
| Years of Schooling | Population-based | 1 | <0.001 | 0.699 | -0.100 (-0.126, -0.073) | 4.10x10^-7^ |
| Intelligence | Population-based | 3 | <0.001 | 0.628 | -0.101 (-0.138, -0.063) | 4.91x10-5 |
| Neuroticism | Population-based | 5 | <0.001 | 0.762 | 0.118 (-0.007, 0.229) | 0.052 |
| *Population-based: population-level mendelian randomization using independent samples from IEU Open GWAS; WFC population-level: population-level Mendelian randomization using independent samples from the Within Family Consortium data; Within-family in sibling pairs: within-family Mendelian randomization using sibling data.* | | | | | | |

| **eTable 7: Bivariate LD score regression intercept between different cohorts using the phenotype height** | | | | | |
| --- | --- | --- | --- | --- | --- |
|  | DiscovEHR | FinnTwin | GenS | QIMR | UKB |
| DiscovEHR | **-** | -0.0002 (0.005) | 0.003 (0.006) | 0.003 (0.007) | 0.009 (0.007) |
| FinnTwin | **-** | **-** | 0.002 (0.006) | -0.004 (0.007) | 0.009 (0.007) |
| GenS | **-** | **-** | **-** | -0.004 (0.006) | 0.012 (0.007) |
| QIMR | **-** | **-** | **-** | **-** | -0.007 (0.007) |
| UKB | **-** | **-** | **-** | **-** | **-** |
| *DiscovEHR =* MyCode Community Health Initiative at Geisinger*, FinnTwin =*Finnish Twin Cohort *, GenS= Generation Scotland, QIMR = QIMR Berghofer Medical Research Institute, UKB = UK Biobank*  *Bivariate LD score regression intercept is a measure of correlated estimation error that can be caused by related or overlapping samples as well as other possible confounding.* | | | | | |

| **eTable 8: IVW Mendelian randomization results for the association between a genetic liability to ADHD and different health outcomes using population-level data from the Within Family Consortium** | | | | | |
| --- | --- | --- | --- | --- | --- |
|  | N SNPs | Beta (SE) | 95% CI | p-value | Adjusted p-value |
| Outcome | | | | | |
| Age at Menopause | 22 | -0.017 (0.031) | -0.077, 0.044 | 0.587 | 0.684* |
| HDL cholesterol | 22 | 0.002 (0.015) | -0.027, 0.031 | 0.906 | 0.906* |
| CRP | 22 | 0.009 (0.015) | -0.020, 0.038 | 0.539 | 0.684 |
| Menarche | 22 | 0.009 (0.023) | -0.036, 0.055 | 0.692 | 0.738* |
| HbA1c | 22 | 0.012 (0.019) | -0.025, 0.049 | 0.525 | 0.684 |
| Triglycerides | 22 | 0.013 (0.016) | -0.018, 0.044 | 0.400 | 0.641* |
| BMI | 22 | 0.018 (0.016) | -0.014, 0.049 | 0.266 | 0.472 |
| WHR | 22 | 0.023 (0.009) | 0.006, 0.040 | 0.008 | 0.040* |
| LDL cholesterol | 22 | 0.027 (0.013) | 0.003, 0.052 | 0.029 | 0.092 |
| Lifestyle factors | | | | | |
| Cognition | 22 | -0.113 (0.027) | -0.166, -0.061 | <0.001 | <0.001* |
| Educational attainment | 22 | -0.060 (0.011) | -0.081, -0.038 | <0.001 | <0.001* |
| Age at first birth | 22 | -0.029 (0.015) | -0.058, -0.001 | 0.044 | 0.116 |
| Neuroticism | 22 | 0.024 (0.016) | -0.007, 0.055 | 0.136 | 0.273* |
| Alcohol intake frequency | 22 | 0.030 (0.056) | -0.080, 0.139 | 0.599 | 0.684 |
| Depressive symptoms | 22 | 0.031 (0.019) | -0.007, 0.068 | 0.115 | 0.263 |
| Cigarettes smoked per day | 22 | 0.049 (0.019) | 0.012, 0.085 | 0.010 | 0.040* |
| * Results where the associations are consistent with analysis using the largest GWAS data available as an outcome. | | | | | |
