## Supplementary info for "Familial factors inflate the estimated effects of genetic liability to ADHD on behavioural outcomes"

### Research in context

#### Literature search strategy

We searched PubMed and Google Scholar from database inception to 20^th^ July 2026, to identify studies investigating attention-deficit hyperactivity disorder, Mendelian randomization and within-family study designs. Eight relevant studies were identified, however only three used a within-family or trio Mendelian randomization design, and none used a large within-family Mendelian randomization design to investigate the effects of genetic liability to ADHD on health or behavioural outcomes. The eight studies are presented in **eTable 1**.

An example PubMed search string was:
*("Attention Deficit Disorder with Hyperactivity" OR “ADHD” OR "attention deficit hyperactivity disorder")
AND
("Mendelian randomization" OR "Mendelian randomisation" OR “MR”)
AND
("within-family" OR "within family" OR "within-sibship" OR “sibling*” OR “sibship” OR “family-based” OR “trio*” OR “duo*” OR” parent-offspring”)*

### Methods

#### Instrument selection and harmonization

SNPs were harmonized, aligning the genetic association for exposure and outcome on the same effect allele. During harmonization, palindromic SNPs with unavailable or intermediate allele frequencies (0.42-0.58) were considered strand ambiguous and removed. SNPs that were unavailable in the outcome datasets were replaced with proxy SNPs with R^2^ > 0.80. We ensured that SNPs were independent through clumping the harmonized SNPs for LD (R^2^=0.001; 10,000kb).

Post-harmonisation, 22 SNPs were available to investigate the effect of a genetic liability to ADHD on the health/lifestyle outcomes in both the population-based and within-family analyses. There was a small variation in the number of available SNPs for each analysis given that they were not all available in every outcome GWAS. SNP sets were harmonised across population-based and within-family analyses meaning that both designs used identical instruments for each phenotype. We calculated an F-statistic for each SNP to assess the strength of association between the instrument and ADHD^[8]^. The F-statistic is a function of the proportion of variance in the phenotype explained by the genetic variants, the sample size, and the number of instruments, where a larger F-statistic suggests a lower risk of weak-instrument bias^[9]^.

#### Investigation of robustness to inflation

Due to a lack of available family-based data, our primary analysis uses SNP-exposure associations from large population-based GWAS. The Wald-type estimators calculate SNP-Outcome/SNP-Exposure. Therefore, social and environmental factors may influence the denominator, despite being controlled in the numerator. Consequently, if our SNP-exposure estimates are inflated, the family-based Mendelian randomization estimates may still be inconsistent. To examine this, we rescaled the SNP-exposure associations by 1-*s,* where *s* represents the degree of shrinkage, and then recalculated the IVW under different levels of SNP-exposure inflation.

#### Assessment of the Mendelian randomization assumptions using UKB data

Using individual-level data from the UK Biobank (UKB)^[1]^, we investigated whether the genetic variants were associated with maternal smoking status during pregnancy or with own birthweight. This was to check for potential violations of the second Mendelian randomization assumption (the independence assumption) and for horizontal pleiotropy (violations of the exclusion-restriction assumption), respectively. Evidence of an association would suggest residual confounding or that the SNPs influence the outcome through biological pathways other than ADHD liability, which would make the causal estimates inconsistent ^[2, 3]^.

#### Assessment of heterogeneity between population-based and within-family samples

In our primary analysis, we used the largest available population-based GWAS to maximise power; however, systematic differences may exist between individuals in the population-based GWAS and those in the within-sibship meta-analysis GWAS. If present, these differences could introduce heterogeneity in the causal effect estimates, which could then be interpreted as a consequence of correcting for environmental/familial factors. Therefore, we ran a second population-based Mendelian randomization analysis using summary statistics from the Within Family Consortium^[4]^ (i.e. these analyses were not performed within families but rather across individuals/families). Consistent results between the large, independent population-based GWAS and the smaller population-based Within Family Consortium GWAS would suggest the cohorts are broadly comparable and that any observed heterogeneity between the models is unlikely to reflect systematic differences among the included individuals.

#### Standardisation procedure

Effect estimates were converted to standard deviation (SD) units to aid with interpretation and comparability across the Mendelian randomization designs. To achieve this, where necessary, the Mendelian randomization estimates (β and SE) originally expressed in raw outcome units were divided by the population standard deviation of the outcome. Where possible, we based the standard deviation for each phenotype on UKB estimates. When these estimates were unavailable, we obtained them from other large-scale, population-based cohorts. Raw units and population standard deviation used for conversion are presented in **eTable 3.** Mendelian randomization estimates and standard errors were then multiplied by ln(2) (0.693), meaning that estimates are expressed as standard deviation units of the outcome per doubling of the genetically predicted odds of ADHD.

#### Sample overlap in cohorts

We assumed a conservative covariance of zero between the population-based Mendelian randomization estimates and the family-based Mendelian randomization estimates because the GWAS were performed in separate cohorts making substantial sample overlap unlikely. For the phenotype height, we ran a series of bivariate LDSC analyses across five of the cohorts that contribute to the sibship within-family SNP-outcome estimates: The Finnish Twin Cohort, Generation Scotland, UK Biobank, MyCode Community Health Initiative at Geisinger and *QIMR Berghofer Medical Research Institute*. SNP-based bivariate genetic correlations (r_g_) were calculated using the LD score regression software for the siblings^[5]^. The cross-trait genetic covariance intercept is a measure of correlated estimation error that can be caused by related or overlapping samples, as well as other potential confounding factors. Here we used it to infer sample overlap. We utilised pre-computed LD score estimates from the HapMap3 European reference panel for ~1.2 million common SNPs. We found very minimal evidence of sample overlap (cross-trait LDSC intercept ≤0.012; **eTable 7**). This provides reassurance for the use of a conservative covariance of 0 in the Wald test used to investigate heterogeneity between the different Mendelian randomization designs.

#### Discussion of within-family frameworks

Population-based and family-based Mendelian randomization differ in how they control for the offspring’s familial genetic background at the molecular genetic level^[6]^. By using the genetic differences between siblings or conditioning on parental genotypes, within family effect estimates are robust to effects from population stratification, assortative mating and dynastic effects^[7, 8]^.

However, while within-sibship and complete trio analyses exploit the segregation of alleles within families, parent-offspring duo analyses condition on the genotype of one parent. This means that assumptions differ across the different within-family designs and they are also liable to different sources of inconsistency.

##### Population-based Mendelian randomization

Population-based Mendelian randomization uses estimates of the SNP-exposure and SNP-outcome association estimated in samples of unrelated individuals. We assume the following underlying model in the population:

$$X_{i}= \gamma_{0}+\gamma_{1}G_{i}+\gamma_{2}U_{i}+\varepsilon_{i}^{X}$$

(1)

$$Y_{i}= \beta_{0}+\beta_{1}Xi+\beta_{2}G_{i}+\beta_{3}U_{i}+\varepsilon_{i}^{Y}$$

(2)

Where:
$G_{i}$_,_ $X_{i}$ and $Y_{i}$ are the genotype, exposure and outcome for individual i;›
$U_{i}$ is a latent variable that represents all the unmeasured confounders of the exposure-outcome relationship, including ancestry, family socioeconomic position, and parental genotypes acting through the individual’s environment;

$\beta_{1}$: is the causal effect of the exposure on the outcome;

$\gamma_{1}:$ is the effect of the SNP on the exposure;

$\gamma_{2}$and $\beta_{3}$ are the effects of the confounder on the exposure and the outcome respectively;

$\beta_{2}$is the direct effect of the genotype on the outcome not mediated by the exposure;

$\gamma_{0}$ and $\beta_{0}$are the exposure and outcome model intercepts;
and $\varepsilon_{i}^{X}$ and $\varepsilon_{i}^{Y}$ are the error terms for the exposure and outcome.

Substituting (1) into (2) gives the reduced form:

$$Y_{i} = (\beta_{0}+\beta_{1}\gamma_{0}) + (\beta_{1}\gamma_{1}+\beta_{2})G_{i} + (\beta_{1}\gamma_{2}+\beta_{3})U_{i} + \beta_{1}\varepsilon_{i}^{X} + \varepsilon_{i}^{Y}$$

(3)

We regress the exposure and outcome on $G_{i}$ only and define:

$$\delta=\frac{COV\left( G,U \right)}{VAR\left( G \right)}$$

(4)

where COV and VAR refer to population level quantities. Taking the probability limit of both regression coefficients and assuming that the constants and is independent of $G_{i}$ yields:

$$\mathrm{plim}\left( \hat{\gamma}^{pop} \right)=\mathrm{plim}\left( \frac{cov\left( X,G \right)}{var\left( G \right)} \right)=\frac{COV\left( X,G \right)}{VAR\left( G \right)}$$

(5)

$$=\frac{COV\left( \gamma_{0},G \right)+\gamma_{1}VAR\left( G \right)+\gamma_{2}COV\left( G,U \right)+COV\left( G,\varepsilon^{X} \right)}{VAR\left( G \right)}$$

(6)

$$=\gamma_{1}+\gamma_{2}\delta$$

$${\mathrm{plim}(\hat{\beta}}^{pop})=\mathrm{plim}\left( \frac{cov\left( Y,G \right)}{var\left( G \right)} \right)=\frac{COV\left( Y,G \right)}{VAR\left( G \right)}$$

(7)

$$=\frac{COV\left( \beta_{0}+\beta_{1}\gamma_{0},G \right)+\left( \beta_{1}\gamma_{1}+\beta_{2} \right)VAR\left( G \right)+(\beta_{1}\gamma_{2}+\beta_{3})COV\left( G,U \right)+COV\left( G,\beta_{1}\varepsilon^{X} +\varepsilon^{Y} \right)}{VAR\left( G \right)}$$

(8)

$$=\beta_{1}\gamma_{1}+\beta_{2}+(\beta_{1}\gamma_{2}+\beta_{3})\delta$$

where plim denotes the probability limit as $N\to\infty$, cov and var are sample variances and covariances, and $COV$and $VAR$ are population variances and covariances.

The Wald ratio for variant *j* is the ratio of the sample regression coefficients for the SNP-outcome and SNP-exposure associations. Taking the probability limit of this estimator gives:

$$\mathrm{plim}(\hat{\beta}_{j})=\mathrm{plim}\left( \frac{\hat{\beta}^{pop}}{\hat{\gamma}^{pop}} \right)$$

(9)

$$=\frac{\beta_{1}\gamma_{1}+\beta_{2}+(\beta_{1}\gamma_{2}+\beta_{3})\delta}{\gamma_{1}+\gamma_{2}\delta}$$

$$=\beta_{1}+\frac{\beta_{2}+\beta_{3}\delta}{\gamma_{1}+\gamma_{2}\delta}$$

Therefore, the Wald ratio is a consistent estimator of the causal effect of the exposure on the outcome in the absence of horizontal pleiotropy ($\beta_{2}=0 )$, and when either the genetic variant is uncorrelated with the unmeasured confounders ($COV(G,U)=0)$, or the unmeasured confounder does not affect the outcome independent of the exposure ($\beta_{3}=0)$.

Hence, Mendelian randomization requires the independence assumption (i.e.
$COV\left( G,U \right)=$*0*), that genetic instruments are independent of unmeasured confounders. Population stratification, assortative mating and dynastic effects could violate this assumption by inducing associations between the genetic instruments and the unmeasured causes of the outcome. This would therefore make the Wald estimator inconsistent for the causal effect when Mendelian randomization is performed on population-based samples of unrelated individuals.

##### Within-family Mendelian randomization using sibships

Siblings from the same family share the same parents, the same ancestry and often aspects of the same environment. Within sibship tests of genetic association capitalize on the random segregation of alleles from parents to their children and are robust to confounding from population stratification, assortative mating and dynastic effects.

Consider the following family-fixed-effects model, as specified in Brumpton *et al*. (2020) ^[9]^. For sibling *i* in family *k*:

$$X_{k,i}= \gamma_{0}+\gamma_{1}G_{k,i}+\gamma_{2}U_{k,i}+f_{k}^{X}+\varepsilon_{k,i}^{X}$$

(10)

$$Y_{k,i}= \beta_{0}+\beta_{1}X_{k,i}+\beta_{2}G_{k,i}+\beta_{3}U_{k,i}+f_{k}^{Y}+\varepsilon_{k,i}^{Y}$$

(11)

where $f_{k}^{X}$ and $f_{k}^{Y}$ are the family-level terms that capture additive contributions that are constant across siblings within family k, including ancestry, socioeconomic position, and parental genotypes acting through the offspring environment. $U_{k,i}$ denotes confounding that differs between siblings within the same family. All other terms are as in equations (1) and (2) above. Note that $f_{k}^{X}$ and $f_{k}^{Y}$ may correlate with genotype, and hence in the population model can cause confounding.

###### The within transformation

Each family has $n_{k}$ siblings. We can therefore take the mean of the exposure (10) and the outcome (11) across the siblings in that family using the function $\bar{A}_{k}=\frac{1}{n_{k}}\sum_{i=1}^{n_{k}} A_{k,i}$

$$\bar{X}_{k}= \gamma_{0}+\gamma_{1}\bar{G}_{k}+\gamma_{2}\bar{U}_{k}+f_{k}^{X}+\bar{\varepsilon}_{k}^{X}$$

(12)

$$\bar{Y}_{k}= \beta_{0}+\beta_{1}\bar{X}_{k}+\beta_{2}\bar{G}_{k}+\beta_{3}\bar{U}_{k}+f_{k}^{Y}+\bar{\varepsilon}_{k}^{Y}$$

(13)

In the averages, two terms pass through without being changed: the constants and the family fixed effects. Therefore, when we subtract the family mean from each term, these terms cancel:

$$X_{k,i}-\bar{X}_{k}= \left( \gamma_{0}-\gamma_{0} \right)+\gamma_{1}\left( G_{k,i}-\bar{G}_{k} \right)+\gamma_{2}\left( U_{k,i}-\bar{U}_{k} \right)+\left( f_{k}^{X}-f_{k}^{X} \right)+\left( \varepsilon_{k,i}^{X}-\bar{\varepsilon}_{k}^{X} \right)$$

(14)

$${Y_{k,i}-\bar{Y}}_{k}= \left( \beta_{0}-\beta_{0} \right)+\beta_{1}\left( X_{k,i}-\bar{X}_{k} \right)+\beta_{2}\left( G_{k,i}-\bar{G}_{k} \right)+\beta_{3}\left( U_{k,i}-\bar{U}_{k} \right)+\left( f_{k}^{Y}-f_{k}^{Y} \right)+\left( \varepsilon_{k,i}^{Y}-\bar{\varepsilon}_{k}^{Y} \right)$$

(15)

Hence the constants and the family fixed effects drop out of the equations. The remaining variation is entirely within families: between-family differences in genotype, in shared environment and in ancestry contribute nothing to the estimate. Note that this transformation requires only sibling data, parental genotypes are not needed.

Let $\Delta A_{k,i}=A_{k,i}-\bar{A}_{k}$:

$${\Delta X}_{k,i}= \gamma_{1}\Delta G_{k,i}+\gamma_{2}{\Delta U}_{k,i}+{\Delta\varepsilon}_{k,i}^{X}$$

(16)

$${\Delta Y}_{k,i}= \beta_{1}{\Delta X}_{k,i}+\beta_{2}{\Delta G}_{k,i}+\beta_{3}\Delta U_{k,i}+{\Delta\varepsilon}_{k,i}^{Y}$$

(17)

The confounder $U_{k,i}$may still be correlated with the differenced genotype. Define the within-family analogue of $\delta$ in (4):

$$\delta_{within}=\frac{COV\left( \Delta G_{k},{\Delta U}_{k} \right)}{VAR\left( {\Delta G}_{k} \right)}$$

(18)

Taking covariances with $\Delta G_{k,i}$ and dividing by $VAR\left( \Delta G_{k} \right)$, as in (5) and (6), and assuming the differenced error terms are uncorrelated with $\Delta G_{k,i}$ gives

$$\mathrm{plim}(\hat{\gamma}^{within})=\gamma_{1}+\gamma_{2}\delta_{within}$$

(19)

$$\mathrm{plim}(\hat{\beta}^{within})=\beta_{1}\gamma_{1}+\beta_{2}+\left( \beta_{1}\gamma_{2}+\beta_{3} \right)\delta_{within}$$

(20)

The Wald ratio for variant *j* is the ratio of the genetic variant outcome and genetic variant exposure associations:

$$\mathrm{plim}(\hat{\beta}_{within,j})=\mathrm{plim}\left( \frac{\hat{\beta}^{within}}{\hat{\gamma}^{within}} \right)$$

(21)

$$=\frac{\beta_{1}\gamma_{1}+\beta_{2}+\left( \beta_{1}\gamma_{2}+\beta_{3} \right)\delta_{within}}{\gamma_{1}+\gamma_{2}\delta_{within}}$$

$$=\beta_{1}+\frac{\beta_{2}+\beta_{3}\delta_{within}}{\gamma_{1}+\gamma_{2}\delta_{within}}$$

Mendel’s laws imply that each offspring’s genotype is a random independent draw from the parental genotypes. Therefore, anything that is determined before conception and shared by siblings must go into $f_{k}$and there can be no other terms that cause $\Delta G_{k,i}.$ Thus, absent of sibling indirect effects, any correlates with $\Delta G_{k,i}$ must be its consequences. Hence, $\delta_{within}=0$. This implies:

$${plim(\hat{\beta}}_{within,j})=\frac{\beta_{1}\gamma_{1}+\beta_{2}}{\gamma_{1}}=\beta_{1}+\frac{\beta_{2}}{\gamma_{1}}$$

(22)

Thus, the within estimate reflects only the direct effects $\beta_{1}$, plus the indirect horizontal pleiotropic effects, $\frac{\beta_{2}}{\gamma_{1}}.$

Equivalently, you can obtain the same estimates by including a dummy variable for each family. Note that this is distinct from taking pairwise differences between siblings, which coincides with (14) and (15) only when each family contributes two siblings. For larger sibships, equivalence depends on how the pairwise differences are weighted. For example, using all pairwise differences without weighting gives larger families additional weight when sibships sizes vary.

##### Within-family Parent-offspring trio Mendelian randomization

When running a mother-father-offspring within-family Mendelian randomization, the offspring SNP-outcome association is conditional on the maternal and paternal genotypes. This controls for the confounding arising from genetic influences on the offspring’s environment and from between-family differences associated with maternal and paternal genotypes.

$$X_{i}=\gamma_{0}+\gamma_{1}G_{i}+\gamma_{m}G_{m(i)}+\gamma_{f}G_{f(i)}+\gamma_{2}U_{i}+\varepsilon_{i}^{X}$$

(23)

$$Y_{i}=\beta_{0}+\beta_{1}X_{i}+\beta_{2}G_{i}+\beta_{m}G_{m(i)}+\beta_{f}G_{f(i)}+\beta_{3}U_{i}+\varepsilon_{i}^{Y}$$

(24)

The indirect, dynastic effects from the mother and father are indicated by $\gamma_{m},$ $\gamma_{f}$, $\beta_{m}$ and $\beta_{f}$. The confounder $U_{i}$ now indicates the effects of all factors not attributable to parental genotype or caused by the offspring’s genotype $G_{i}$. Given that, conditional on both parental genotypes, the offspring genotype is determined solely by which alleles were transmitted, and transmission is random, $G_{i}$ is independent of anything determined before conception. As such we can assume that $COV(U,G\left| G_{m}, G_{f} \right)=0$.

Substituting (23) into (24) and collecting gives:

$$Y_{i}=\left( \beta_{0}+\beta_{1} \gamma_{0} \right)+\Gamma_{1}G_{i}+\Gamma_{m}G_{m(i)}+\Gamma_{f}G_{f(i)}+\left( {\beta_{1}\gamma}_{2}+\beta_{3} \right)U_{i}+{\beta_{1}\varepsilon}_{i}^{X}+\varepsilon_{i}^{Y}$$

(25)

Where $\Gamma_{1}=\beta_{1}\gamma_{1}+\beta_{2}$, $\Gamma_{m}=\beta_{1}\gamma_{m}+\beta_{m}$, and $\Gamma_{f}=\beta_{1}\gamma_{f}+\beta_{f}$. As $COV(U,G\left| G_{m}, G_{f} \right)=0$, the coefficient on $G_{i}$ in the multivariable regression of $X_{i}$ on $G_{i}$, $G_{m(i)}$, and $G_{f(i)}$ identifies $\gamma_{1}$, and the corresponding coefficient in the regression of $Y_{i}$ identifies $\Gamma_{1}$. In the notation of (4), the trio design sets $\delta=0$.

The analogous Wald estimator for the trios model is therefore asymptotically identical to the sibling estimand:

$${\mathrm{plim}(\hat{\beta}}^{trio})=\frac{\Gamma_{1}}{\gamma_{1}}=\beta_{1}+\frac{\beta_{2}}{\gamma_{1}}$$

(26)

Thus, asymptotically, the trio estimate reflects the direct causal effect of interest $\beta_{1}$, plus an additional term due to horizontal pleiotropy, $\frac{\beta_{2}}{\gamma_{1}}$.

##### Within-family Parent-offspring duo Mendelian randomization

For many samples, maternal and offspring genotypes are available, but paternal genotype is not. You can estimate the effect of an exposure using these samples, but you cannot control for the indirect effect of the paternal genotype. These effects may confound the estimated exposure-outcome association which, in econometrics, is referred to as an example of omitted variable bias. Since $U_{i}$ excludes factors attributable to parental genotype, it remains uncorrelated with $G_{i}$ given $G_{m(i)}$, and the confounding in the duo design operates through the omitted $G_{f(i)}$.

###### Deriving the inconsistency in the estimator

We decompose the omitted variable into a part that the included regressors can explain and a part that they cannot:

$$G_{f(i)} = \delta_{0} + \delta_{f}G_{i} + \delta_{m}G_{m(i)} + r_{i}$$

(27)

By construction $\mathrm{COV}(r,G) = \mathrm{COV}(r,G_{m}) = 0$. Substituting (27) into the reduced form (25), and writing $c_{0}$ for the intercept $(\beta_{0}+\beta_{1}\gamma_{0})$ and $W_{i}=(\beta_{1}\gamma_{2}+\beta_{3})U_{i} + \beta_{1}\varepsilon_{i}^{X} + \varepsilon_{i}^{Y}$, and collecting terms in the two observed genotypes:

$$Y_{i} = (c_{0}+\Gamma_{f}\delta_{0}) + (\Gamma_{1}+\Gamma_{f}\delta_{f})G_{i} + (\Gamma_{m}+\Gamma_{f}\delta_{m})G_{m(i)} + (\Gamma_{f}r_{i}+W_{i})$$

(28)

This is the regression of $Y_{i}$ on $G_{i}$ and $G_{m(i)}$, and it is the population regression, if its error term is uncorrelated with both regressors. This holds here by construction (27). The coefficients in (28) are therefore:

$$\mathrm{plim}(\hat{\Gamma}_{1}^{\mathrm{duo}} )= \Gamma_{1} + \delta_{f}\Gamma_{f}$$

(29)

$${\mathrm{plim}(\hat{\Gamma}}_{m}^{\mathrm{duo}}) = \Gamma_{m} + \delta_{m}\Gamma_{f}$$

(30)

Thus, the reduced-form offspring and maternal coefficients are inconsistent whenever there is a paternal reduced-form effect, $\Gamma_{f}\neq0$.

###### Evaluating the projection coefficients

We can decompose each genotype into its constituent alleles. Let $G_{i} = A_{m} + A_{f}$, where $A_{m}$ and $A_{f}$ are the maternally and paternally transmitted alleles; let $G_{m(i)} = A_{m} + A_{m}^{'}$ and $G_{f(i)} = A_{f} + A_{f}^{'}$, where the primed terms are the non-transmitted alleles. Under Hardy-Weinberg equilibrium and random mating, the four alleles are independent, each with variance $s = p(1-p)$, where $p$ is the effect allele frequency. Hence $\mathrm{VAR}(G) = \mathrm{VAR}(G_{m}) = \mathrm{VAR}(G_{f(i)}) = 2s$, $\mathrm{COV}(G,G_{m}) = \mathrm{VAR}(A_{m}) = s$, $\mathrm{COV}(G,G_{f}) = \mathrm{VAR}(A_{f}) = s$, and $\mathrm{COV}(G_{m},G_{f}) = 0$, the last following from random mating.

The normal equations for the projection in (27) are therefore:

$$2s\delta_{f} + s\delta_{m} = s$$

(31)

$$s\delta_{f} + 2s\delta_{m} = 0$$

(32)

which solve to:

$$\delta_{f} = \frac{2}{3}, \delta_{m} = -\frac{1}{3}$$

(33)

The value $\delta_{f} = \frac{2}{3}$ depends on random mating only through $\mathrm{COV}(G_{m},G_{f}) =0$. Under assortative mating, this covariance is positive, the terms in (31) and (32) change, and $\delta_{f}$ departs from 2/3. Intuitively, $G_{i}$ carries information about $G_{f(i)}$ through the transmitted allele $A_{f}$, but does so noisily because $A_{m}$ is also present. Conditioning on $G_{m(i)}$ removes part of that noise, though not all of it, since $G_{m(i)}$ also contains the non-transmitted allele $A_{m}^{'}$.

###### The duo Wald ratio

The same omission applies to the exposure model, giving:

$${\mathrm{plim}(\hat{\gamma}}_{1}^{\mathrm{duo}}) = \gamma_{1} + \frac{2}{3}\gamma_{f}$$

(34)

Forming the Wald ratio:

$${\mathrm{plim}(\hat{\beta}}^{\mathrm{duo}}) = \frac{\Gamma_{1}+\frac{2}{3}\Gamma_{f}}{\gamma_{1}+\frac{2}{3}\gamma_{f}}$$

(35)

$$= \beta_{1} + \frac{\beta_{2}+\frac{2}{3}\beta_{f}}{\gamma_{1}+\frac{2}{3}\gamma_{f}}$$

Two features of (35) are worth noting. First, the $\beta_{1}\gamma_{f}$ term cancels, for the same reason that the $\beta_{1}\gamma_{2}\delta$ term cancelled in (9): paternal genotype acting on the outcome through the offspring's exposure forms part of the causal effect being estimated, and is not confounding. What remains is $\frac{2}{3}\beta_{f}$, the part of the paternal effect that reaches the outcome without passing through the exposure.

Compare the three estimators side by side:

| **Probability limits of the Wald ratio** | |  |
| --- | --- | --- |
| Population (9) | $\beta_{1} + \frac{\beta_{2}+\beta_{3}\delta}{\gamma_{1}+\gamma_{2}\delta}$ | Confounding and horizontal pleiotropy when present may produce inconsistency |
| Duo (35) | $\beta_{1} + \frac{\beta_{2}+\frac{2}{3}\beta_{f}}{\gamma_{1}+\frac{2}{3}\gamma_{f}}$ | Removes effects from the included parent’s contribution but can retain effects from the omitted parent which may produce inconsistency |
| Sibship / trio (22 and 26) | $\beta_{1} + \frac{\beta_{2}}{\gamma_{1}}$ | Removes familial contribution under their respective assumptions, but neither automatically removes horizontal pleiotropy through $\frac{\beta_{2}}{\gamma_{1}}$ |

The duo design removes the maternal indirect effect entirely but retains two-thirds of the paternal one, and reduces exactly to the trio estimand when $\beta_{f} = \gamma_{f} = 0$. It does not, however, uniformly dominate the population-based design. Omitting both parental genotypes gives each parent a weight of $\frac{COV\left( G_{m},G \right)}{VAR\left( G \right)}=\frac{1}{2}$, so conditioning on the mother raises the weight on the paternal effect from 1/2 to 2/3: partialling out $G_{m(i)}$removes the $A_{m}$ component of $G_{i}$and makes it a sharper proxy for $A_{f}$, and hence for $G_{f(i)}$. Where maternal indirect effects are small relative to paternal ones, the duo estimate can therefore carry more paternal contamination than the population-based estimate, though it will usually improve on it because the maternal term is eliminated. The case of a present paternal genotype and missing maternal genotype is analogous.

### Results

#### Assessment of heterogeneity between population-based and within-family samples

Results from the population-level Mendelian randomization using the smaller within family consortium data were mostly consistent with the result from our main analysis.

Namely, we found that the population-based Mendelian randomization using Within Family Consortium summary statistics suggests that doubling a person’s genetic liability to ADHD the number of cigarettes they smoked per day by 0.049 (95%CI: 0.012, 0.085) , reduced educational attainment by 0.060 (95%CI:-0.081, -0.038) and reduced cognitive performance by 0.113 (95%CI: -0.166, -0.061).

Population-based Mendelian randomization using the Within Family Consortium summary statistics also found that doubling a person’s genetic liability to ADHD increased their waist-to-hip ratio by 0.023 (95%CI:0.006, 0.040). All results for the IVW Mendelian randomization analysis are presented in **eTable 8.** Given the much smaller sample size, a lack of an effect where one was previously observed, such as alcohol consumption, is likely a lack of power.

We observed evidence of heterogeneity among the instrumental variables for the phenotype BMI (Q=56.77; p-value=<0.001) and HDL-cholesterol (Q=103.11; p-value=0.037). Results are presented in **eTable 5.** When evaluating pleiotropy using MR-PRESSO, horizontal pleiotropic outliers were detected for a small number of associations. However, like in the main analysis, removal only had a minor effect on the strength of the previously reported relationships. Identified outliers and pleiotropy-corrected associations measured by MR-PRESSO are presented in **eTable 6.**
