## Supplementary material for "Familial factors inflate the estimated effects of genetic liability to ADHD on behavioural outcomes": MR Strobe checklist

**STROBE-MR checklist of recommended items to address in reports of Mendelian randomization studies**^1^ ^2^

| **Item No.** | **Section** | **Checklist item** | **Page No.** | **Relevant text from manuscript** |
| --- | --- | --- | --- | --- |
| 1 | **TITLE and ABSTRACT** | Indicate Mendelian randomization (MR) as the study’s design in the title and/or the abstract if that is a main purpose of the study | 3 | We used up to 24 genome-wide significant variants associated with ADHD (38,691 cases, 186,843 controls) to estimate effects of genetic liability to ADHD on 16 health and behavioural outcomes. We compared population-based estimates with family-based estimates from within-sibship analyses of up to 164,220 siblings. We tested for heterogeneity between designs, and used parent–offspring duos and trios. |
|  | **INTRODUCTION** |  |  |  |
| 2 | **Background** | Explain the scientific background and rationale for the reported study. What is the exposure? Is a potential causal relationship between exposure and outcome plausible? Justify why MR is a helpful method to address the study question | 5-6 | e.g. However, the majority, if not all, of the Mendelian randomization studies to date investigating the effects of ADHD on health have used samples of unrelated individuals, which we will refer to as population-based Mendelian randomization. Given that Mendelian segregation randomizes genetic variants *within families*^[25]^, population-based Mendelian randomization estimates may be inconsistent if genetic variants associated with ADHD are not randomly distributed across the population^[26-29]^ (i.e. there is uncontrolled confounding of the genetic variant -outcome association), as this would violate the independence assumption.  **…**  Further rational in manuscript |
| 3 | **Objectives** | State specific objectives clearly, including pre-specified causal hypotheses (if any). State that MR is a method that, under specific assumptions, intends to estimate causal effects | 6 | In the present study, we compared two-sample population-based and within-family Mendelian randomization estimates of the effect of genetic liability to ADHD on health, behavioural and social outcomes. Given that within-family designs separate a person's own genetic liability from the familial and environmental factors that confound conventional genetic studies, this tests whether the outcomes associated with ADHD reflect the individual's own ADHD. To our knowledge, this is the largest molecular genetic family-based study of ADHD to date. |
|  | **METHODS** |  |  |  |
| 4 | **Study design and data sources** | Present key elements of the study design early in the article. Consider including a table listing sources of data for all phases of the study. For each data source contributing to the analysis, describe the following: | 9, 10 | Data source presented in Table 3 |
|  | a) | Setting: Describe the study design and the underlying population, if possible. Describe the setting, locations, and relevant dates, including periods of recruitment, exposure, follow-up, and data collection, when available. | 10, 11 |  |
|  | b) | Participants: Give the eligibility criteria, and the sources and methods of selection of participants. Report the sample size, and whether any power or sample size calculations were carried out prior to the main analysis | 9-11 | Supplementary table 3, |
|  | c) | Describe measurement, quality control and selection of genetic variants | 9, 10 | e.g. Genome-wide significant single-nucleotide polymorphisms (SNPs) (P<5.0×10^-8^) were used as instruments for ADHD and identified from summary-level population-based data from a European GWAS meta-analysis (38,691 individuals with ADHD and 186,843 controls). |
|  | d) | For each exposure, outcome, and other relevant variables, describe methods of assessment and diagnostic criteria for diseases | 10 | Further information presented in the supplement |
|  | e) | Provide details of ethics committee approval and participant informed consent, if relevant | 20 | UK Biobank has approval from the North West Multi-centre Research Ethics Committee (MREC) as a Research Tissue Bank (RTB) approval. |
| 5 | **Assumptions** | Explicitly state the three core IV assumptions for the main analysis (relevance, independence and exclusion restriction) as well assumptions for any additional or sensitivity analysis | 5 | Assumption 1 (Relevance): The instrument (G) is robustly associated with the exposure (X).  Assumption 2 (Independence): There are no uncontrolled confounders of the association between the instruments and the outcome, i.e. the instrument does not have a common cause with the outcome.  Assumption 3 (Exclusion-restriction): The instrument influences the outcome (Y) only through its effect on the exposure. |
| 6 | **Statistical methods: main analysis** | Describe statistical methods and statistics used | 10, 11 |  |
|  | a) | Describe how quantitative variables were handled in the analyses (i.e., scale, units, model) | 11 | e.g. Outcomes were standardised using the population standard deviation. Mendelian randomization effect estimates and standard errors were multiplied by ln(2) (0.693), meaning that estimates are expressed as standard deviation units of the outcome per doubling of the genetically predicted odds of ADHD.  Further information presented in the supplement |
|  | b) | Describe how genetic variants were handled in the analyses and, if applicable, how their weights were selected | 10 | Further information presented in the supplement |
|  | c) | Describe the MR estimator (e.g. two-stage least squares, Wald ratio) and related statistics. Detail the included covariates and, in case of two-sample MR, whether the same covariate set was used for adjustment in the two samples | 10,11 |  |
|  | d) | Explain how missing data were addressed |  | N/A |
|  | e) | If applicable, indicate how multiple testing was addressed | 11 |  |
| 7 | **Assessment of assumptions** | Describe any methods or prior knowledge used to assess the assumptions or justify their validity | 9, 12 |  |
| 8 | **Sensitivity analyses and additional analyses** | Describe any sensitivity analyses or additional analyses performed (e.g. comparison of effect estimates from different approaches, independent replication, bias analytic techniques, validation of instruments, simulations) | 11-13, 16 |  |
| 9 | **Software and pre-registration** |  |  |  |
|  | a) | Name statistical software and package(s), including version and settings used | 20 | Code for data management and statistical analysis is available at: <https://github.com/daisycrick>.  SOFTWARE: R version 4.5.2, Plink1.9; data manipulation: dplyr, tidyverse ; Packages: TwoSampleMR, MendelianRandomization, MR-PRESSO; Visualisation: forestplotter, ggplot2.  Study protocol was not pre-registered. |
|  | b) | State whether the study protocol and details were pre-registered (as well as when and where) |  |  |
|  | **RESULTS** |  |  |  |
| 10 | **Descriptive data** |  |  |  |
|  | a) | Report the numbers of individuals at each stage of included studies and reasons for exclusion. Consider use of a flow diagram |  | Supplementary table 3 |
|  | b) | Report summary statistics for phenotypic exposure(s), outcome(s), and other relevant variables (e.g. means, SDs, proportions) |  |  |
|  | c) | If the data sources include meta-analyses of previous studies, provide the assessments of heterogeneity across these studies |  |  |
|  | d) | For two-sample MR:  i.  Provide justification of the similarity of the genetic variant-exposure associations between the exposure and outcome samples  ii.  Provide information on the number of individuals who overlap between the exposure and outcome studies | 11 |  |
| 11 | **Main results** |  |  |  |
|  | a) | Report the associations between genetic variant and exposure, and between genetic variant and outcome, preferably on an interpretable scale | 13-15 |  |
|  | b) | Report MR estimates of the relationship between exposure and outcome, and the measures of uncertainty from the MR analysis, on an interpretable scale, such as odds ratio or relative risk per SD difference | 13-15 |  |
|  | c) | If relevant, consider translating estimates of relative risk into absolute risk for a meaningful time period |  |  |
|  | d) | Consider plots to visualize results (e.g. forest plot, scatterplot of associations between genetic variants and outcome versus between genetic variants and exposure) | 15 |  |
| 12 | **Assessment of assumptions** |  |  |  |
|  | a) | Report the assessment of the validity of the assumptions | 12, 13, 17 | Further information presented in the supplement |
|  | b) | Report any additional statistics (e.g., assessments of heterogeneity across genetic variants, such as *I^2^*, Q statistic or E-value) | 12, 13, 17 | Further information presented in the supplement |
| 13 | **Sensitivity analyses and additional analyses** |  |  |  |
|  | a) | Report any sensitivity analyses to assess the robustness of the main results to violations of the assumptions | 17 | Further information presented in the supplement |
|  | b) | Report results from other sensitivity analyses or additional analyses | 12, 13, 17 | Further information presented in the supplement |
|  | c) | Report any assessment of direction of causal relationship (e.g., bidirectional MR) | 13-14 |  |
|  | d) | When relevant, report and compare with estimates from non-MR analyses |  |  |
|  | e) | Consider additional plots to visualize results (e.g., leave-one-out analyses) |  |  |
|  | **DISCUSSION** |  |  |  |
| 14 | **Key results** | Summarize key results with reference to study objectives | 17, 18 | We used data from 38,691 people with ADHD, 186,843 controls and up to 164,220 siblings to compare conventional population-based Mendelian randomization estimates of the effects of genetic liability to ADHD on health, behavioural and social outcomes with family-based estimates. Both designs aim to estimate the causal effect of an individual's genetic liability to ADHD, but population-based estimates are susceptible to confounding by the familial and social environment in which ADHD arises, whereas family-based estimates isolate the effect of an individual's own liability and are robust to population stratification, assortative mating and dynastic effects (outlined in **Box 1** and **Figure 2**)^[25, 52, 53]^. Population-based estimates reproduced results across the observational and Mendelian randomization literature^[21, 22, 24]^, whereas in family-based analyses, most effects attenuated towards the null, suggesting the population-based estimates for depressive symptoms and age at first child were inconsistent. |
| 15 | **Limitations** | Discuss limitations of the study, taking into account the validity of the IV assumptions, other sources of potential bias, and imprecision. Discuss both direction and magnitude of any potential bias and any efforts to address them | 18 | Our study also has limitations. The SNP–exposure associations were derived from a population-based GWAS and could be influenced by familial factors; however, estimates were essentially unchanged across a wide range of shrinkage parameters, suggesting that inflation of the SNP–ADHD association is unlikely to affect our conclusions. Family-based Mendelian randomization is inherently less precise than population-based analysis, because it uses only within-family variation and genotyped relatives remain scarce^[26, 27]^; this likely explains why we could not detect heterogeneity for some outcomes. Our findings highlight the need for larger family-based GWAS and more family-based non-European samples^[25, 52, 53, 56]^. We also did not use Steiger filtering to assess effect direction, as its validity depends on comparable measurement properties and error across the exposure and outcome. In our setting, the exposure is binary and estimated in a separate sample compared to the outcome, which makes such assumptions unlikely. |
| 16 | **Interpretation** |  |  |  |
|  | a) | Meaning: Give a cautious overall interpretation of results in the context of their limitations and in comparison with other studies | 18,19 | e.g. In conclusion, the associations between genetic liability to ADHD and behavioural and social outcomes reported in population-based studies appear, at least in part, to reflect the family environments in which ADHD arises rather than the direct effect of an individual's own liability. These findings suggest that some of the burden attributed to an individual's ADHD may instead reflect familial and social contexts, and that testing causal claims about the individual-level effects of ADHD liability requires familial data. Population-based Mendelian randomization remains valuable, but family-based designs are among the few approaches that can separate direct effects from familial confounding. Progress will depend on assembling much larger samples of genotyped relatives. |
|  | b) | Mechanism: Discuss underlying biological mechanisms that could drive a potential causal relationship between the investigated exposure and the outcome, and whether the gene-environment equivalence assumption is reasonable. Use causal language carefully, clarifying that IV estimates may provide causal effects only under certain assumptions | 17, 18 |  |
|  | c) | Clinical relevance: Discuss whether the results have clinical or public policy relevance, and to what extent they inform effect sizes of possible interventions | 17-18 | e.g. If part of the behavioural and social burden associated with ADHD reflects the family and environmental context in which ADHD arises, rather than the individual's own liability, then interventions directed solely at the individual, including pharmacological treatment, may not reduce these outcomes as much as observational and population-based genetic estimates imply. The same distinction has implications for burden-of-disease and health-economic models, which typically attribute these associations causally to ADHD and may overstate the fraction modifiable by treating the individual. Clinical and policy conclusions should be drawn from bodies of evidence, not from single studies^[54]^; future research should rigorously test whether these implications translate into effective interventions. |
| 17 | **Generalizability** | Discuss the generalizability of the study results (a) to other populations, (b) across other exposure periods/timings, and (c) across other levels of exposure | 18 |  |
|  | **OTHER INFORMATION** |  |  |  |
| 18 | **Funding** | Describe sources of funding and the role of funders in the present study and, if applicable, sources of funding for the databases and original study or studies on which the present study is based | 19 | NMD is supported by the Norwegian Research Council 295989, the Medical Research Council (MRC) MR/V002147/1 and UKRI1510: the National Institute for Mental Health (NIMH) MH130448, and the [Division of Psychiatry](https://www.ucl.ac.uk/psychiatry/division-psychiatry) (https://www.ucl.ac.uk/brain-sciences/psychiatry). SEM is supported by an Australian National Health and Medical Research Council Investigator Grant (2025674). DME and this research is supported by an Australian National Health and Medical Research Council Investigator Grant (2017942). |
| 19 | **Data and data sharing** | Provide the data used to perform all analyses or report where and how the data can be accessed, and reference these sources in the article. Provide the statistical code needed to reproduce the results in the article, or report whether the code is publicly accessible and if so, where | 19,20 | Code for data management and statistical analysis is available at: <https://github.com/daisycrick>. |
| 20 | **Conflicts of Interest** | All authors should declare all potential conflicts of interest | 19 | None declared. |

This checklist is copyrighted by the Equator Network under the Creative Commons Attribution 3.0 Unported (CC BY 3.0) license.

1.	Skrivankova VW, Richmond RC, Woolf BAR, Yarmolinsky J, Davies NM, Swanson SA, et al. Strengthening the Reporting of Observational Studies in Epidemiology using Mendelian Randomization (STROBE-MR) Statement. JAMA. 2021;under review.

2.	Skrivankova VW, Richmond RC, Woolf BAR, Davies NM, Swanson SA, VanderWeele TJ, et al. Strengthening the Reporting of Observational Studies in Epidemiology using Mendelian Randomisation (STROBE-MR): Explanation and Elaboration. BMJ. 2021;375:n2233.
